# Assessing the feasibility of large-scale digital sensing for depression and anxiety: The Digital Mental Health Study

**DOI:** 10.1101/2025.10.01.25337105

**Authors:** Christopher S Douglas, Eliza Congdon, Crane Huang, Darsol Seok, Zachary D Cohen, Samir Akre, Veronica Tozzo, Feiyang Huang, Danielle Ramo-Larios, Raphe A Bernier, Jonathan Flint, Arash Naeim, Brunilda Balliu, Alex AT Bui, Marian Stewart Bartlett, Michelle G Craske, Nelson B Freimer

**Author notes:** Equal contribution (Christopher S Douglas, Eliza Congdon, Crane Huang, Darsol Seok, Marian Stewart Bartlett, Michelle G Craske, Nelson B Freimer).

## Abstract

Data passively obtained from smartphones and wearables can provide nearly continuous objective information that enables quantification of states and traits across broad physiological, behavioral, and emotional domains impacted in mental health conditions, including depression and anxiety. Widespread application of such digital phenotyping could transform the assessment of depression and anxiety in research and clinical care, but the field has lacked well-powered longitudinal studies demonstrating the utility of this approach. This paper describes the design and implementation of the Digital Mental Health Study (DMHS), which collected up to 12 months of sensor data from iPhone and Apple Watch in more than 4,000 participants, a sample diverse by age, sex at birth, ethnicity, and depression symptom severity. To enable the use of these digital phenotypes to assay the complexity and heterogeneity of depression and anxiety, we designed a protocol of periodic self-report and interview-based scales optimized to assess elements of depression, anxiety, and perceived stress as broadly as possible while minimizing participant measurement burden. We report here the strategies used to recruit and enroll the DMHS sample, the process employed to develop study methods and protocols, and initial findings describing longitudinal symptom trajectories and demonstrating high participant engagement and adherence over a 12-month period.

## Introduction

Depression and anxiety are the most common mental health conditions.^1^ They are among the leading contributors to the global burden of disease, affecting several hundred million people worldwide. ^2^ Reducing this burden will require advances in understanding their underlying causes, developing more effective treatments, and more accurately predicting individual patient outcomes.^3,4,5,6^

Current classification frameworks for depression and anxiety largely ignore their heterogeneity, grouping individuals experiencing symptoms into discrete diagnostic categories. For example, a diagnosis of Major Depressive Disorder (MDD) represents more than 200 different possible combinations of symptoms.^7,8^ Assessment of depression and anxiety most commonly is based on elicitation of snapshots of symptoms through clinical interviews or self-report scales. These evaluations are subjective, susceptible to inaccurate recall and other biases, and inadequately capture core elements of these conditions (e.g., those relating to physiology or behavior) or their dynamic nature. ^9^

The ubiquity of smartphones and wearable devices provides an opportunity to transform the classification and assessment of depression and anxiety, through the continuous collection of multimodal data from their sensors to measure features of physiology, behavior, and emotional states that are associated with these conditions.^10^ Such digital phenotyping offers the potential to objectively characterize consenting individuals, on an immense scale. However, despite this potential, the utility of digital phenotyping as a tool for mental health research and clinical care has not yet been convincingly demonstrated.^11^

Although previous research has shown correlations between depression and anxiety symptoms and digital markers (such as levels of activity, sleep disruption, and social communication patterns), conclusions from such studies have been limited by small, demographically homogenous samples, and data collection over too short a duration for longitudinal analyses.^12^ Furthermore, most studies to date have not attempted to integrate data from smartphones with those obtainable from wearables and have analyzed only a fraction of the data streams available on modern devices. ^13^ Additionally, inconsistent participant engagement with, and adherence to longitudinal protocols in digital mental health research has both complicated data interpretation and raised concerns about the feasibility of deploying these tools in clinical settings.^14^

To address these limitations, we designed and implemented the Digital Mental Health Study (DMHS), a prospective investigation to identify the relationship between self-reported depression and anxiety symptoms and digital phenotypes obtained from iPhone and Apple Watch. Prior to conducting the main DMHS study we conducted two small pilot studies to assess feasibility both in terms of participant burden and operational workflows.

In the main DMHS study, we recruited more than 4,000 participants who provided up to 12 months of nearly continuous digital data. The sample was diverse by ethnicity, age, sex at birth, and symptom severity. We conducted digital phenotyping by integrating data streams from iPhone with those from Apple Watch, thus obtaining information on a much broader set of domains across physiology, behavior, and emotional state than would be available through other consumer-grade platforms. To determine the utility of these digital phenotypes for assaying the complexity and heterogeneity of depression and anxiety, we designed a protocol of periodic self-report and interview-based scales. The protocol was optimized to assess elements of depression and anxiety as broadly as possible without being too burdensome for participants. Additionally, because both perceived stress and the experience of stressful events are theorized to be risk factors for both depression and anxiety,^15^ the DMHS incorporated scales measuring these variables, with the aim of analyzing their relation to both symptom assessments and digital phenotypes.

This paper describes the foundational work of the DMHS, including the recruitment strategies used to enroll the sample, the process employed to develop the study methods and protocol, and initial findings demonstrating high participant engagement and adherence over a 12-month period. This information establishes the feasibility of our digital phenotyping approach for conducting large-scale mental health research studies and identifies the elements of depression and anxiety that we aim to illuminate in future analyses of the DMHS data. We suggest how these analyses may advance the development of a novel data-driven framework for unraveling the complexity of these conditions, and of mental health disorders more broadly.

## Methods

The methods described below are for the main (12 months) DMHS. The DMHS protocol was informed by insights from two pilot studies (DMHS Pilot 1 [DMHS-P1] and DMHS Pilot 2 [DMHS-P2]; see *Supplementary Material*).

### Participant Recruitment and Enrollment

Participants were recruited from two sources: patients receiving care within the University of California, Los Angeles (UCLA) Health system and students enrolled at UCLA. Recruitment employed a multifaceted approach, including targeted messages through the UCLA Health patient portal (allowing for stratification based on demographics and clinic affiliation, with a focus on increasing enrollment of peripartum individuals); automated telephone recruitment to UCLA Health System patients; and communications via the UCLA Student Registrar’s office, departmental email lists, campus organizations, social media, email newsletters, and a prospective participant registry. All materials directed individuals to a study website providing details on eligibility, time commitment, and compensation. Prospective participants initiated the process of screening for eligibility via the website. Eligible individuals were presented with an online informed consent form to review and sign electronically. Prospective participants could additionally contact our study team by email or phone prior to initiating online screening, prior to enrolling, or at any time during study participation.

The study aimed for a final sample of 3000 participants completing the study and meeting minimal protocol adherence. To account for anticipated attrition, we initially set an overall recruitment target of 4200 participants (an estimated retention rate of 72%). After the study kicked off, we increased this target to 4500 participants, adding an additional 300 participants to further increase the likelihood of achieving a sample of at least 3000 study completers. Recruitment was stratified by depression symptom severity, as measured by the Patient Health Questionnaire-8 [PHQ-8] ^16^ administered during online screening. Target composition was 30% with minimal or no depressive symptoms (PHQ-8 <5), 30% mild (PHQ-8 5-9), 20% moderate (PHQ-8 10-14), and 20% moderate-severe or severe symptoms (PHQ-8 > 14). The recruitment targets for these “severity bins” were balanced for age, sex at birth, and ethnicity, such that the demographic distribution was consistent across severity.

The target sample composition for study completers was approximately 1000 UCLA students and 2000 UCLA Health System patients. The student cohort was intended to constitute a relatively homogeneous sample for evaluating signal strength and to enable comparison to other digital mental health studies of college students, while the UCLA Health cohort was intended to enable broader analysis of demographic groups and daily routines. We sought to recruit evenly by male and female sex at birth. Additionally, we aimed to include a subsample of peripartum participants as part of either the student or health system cohorts. Within the UCLA Health sample, we aimed for at least 15% representation of each of four age categories. The overall age distribution targets were: 47% aged 18-25, 23% aged 26-45, 15% aged 46-65, and 15% over 65. Regarding self-reported race and ethnicity, we aimed to collect a minimum of 15% in each of four subgroups. With a goal of making the distribution as even as possible within the recruiting time window, the targets doubled the population prevalence of three of the subgroups and capped the most prevalent group. The resulting target distribution was as follows: From among the UCLA Health cohort, we aimed to recruit a sample of at most 45% Non-Hispanic White or Other and at least 21%, 18%, and 16% Asian, Hispanic, and Black, respectively. Among the UCLA students, our target was at most 40% Non-Hispanic White or Other, and at least 30%, 15%, and 15%, Asian, Hispanic, and Black, respectively. Other eligibility requirements are detailed in *Supplementary Table 1*. Briefly, participation required either ownership of an eligible iPhone or agreement to utilize a study-provided loaner iPhone, and willingness to use a study-provided Apple Watch for the duration of the study.

### Participant Safety and Ethical Considerations

All participants gave written electronic informed consent according to the procedures approved by the University of California Los Angeles Institutional Review Board. Additionally, we implemented a protocol, overseen by licensed clinical staff, to follow-up on disclosures of suicidality in clinical interviews and self-report surveys.

The study was conducted in accordance with the Declaration of Helsinki and relevant institutional guidelines and regulations. Ethical approval was obtained from the University of California, Los Angeles Institutional Review Board (DMHS-P1: #20-000995; DMHS-P2: #20-002337; DMHS main study: #21-001558). All participants provided written informed consent electronically prior to participation. A safety protocol, overseen by licensed clinical staff, was implemented to follow up on any disclosures of suicidality in clinical interviews or self-report surveys. Clinical trial number: not applicable.

### Device Provision and Data Collection

To obtain survey and digital sensing data we provided all enrolled participants with an Apple Watch Series 6, and, for those without a compatible device, a loaner iPhone. Participants installed the Apple Research app^17^ on their iPhone to manage collection of sensor and device data, as well as surveys. The app is a platform that enables individuals to securely share data from their devices for research studies, preserving their privacy. Participants were provided with detailed information about all data types to be collected and were required to authorize sharing for each type. They could review and manage their data sharing permissions within the app at any time throughout the study. Data collection spanned 52 weeks, utilizing the provided devices, the Apple Research app on participants’ iPhones, online surveys, and assessment sessions with study staff. Participants were instructed to wear the Apple Watch for at least 20 hours daily, including during sleep, and to charge it for at least two hours daily while it was connected to their iPhone and Wi-Fi.

Intake assessment sessions were conducted upon enrollment. During these sessions, study staff confirmed the iPhone iOS version, paired the Apple Watch, guided participants through Apple Research app enrollment, reviewed the study schedule, reiterated device charging requirements, explained compensation, and emphasized ongoing data monitoring. Check-in assessment sessions, including interviewer-administered rating scales, were conducted remotely or in-person, occurring at approximately weeks 10-13 and 46-49. Participation concluded with an exit assessment. Participants could track completion of active tasks within the Apple Research app itself. In addition, they received quarterly progress reports summarizing average daily mood ratings, via email, and earned compensation on a quarterly basis.

Interviewer-administered rating scales were obtained using Research Electronic Data Capture (REDCap), a platform that was also used to collect a subset of the self-report surveys. Surveys that were collected at high frequency, (as detailed below) were administered using the Apple Research app installed on participants’ iPhones; each survey had its own schedule for task reminders.

### Assessment Protocol

We aimed to implement a protocol that would enable elucidation of the relationship between a broad range of digital phenotypes and traditional assessments of depression and anxiety, obtained by interviewer-administered rating scale and self-report questionnaires. We therefore optimized a protocol to most fully capture the complexity of these conditions, and to collect information about both exposure to stressful events and subjective experience of stress, without creating an unacceptable burden for participants. The design of the symptom assessment protocol followed a three-step process (*Figure 1*) with the goal of comprehensively evaluating core constructs while reducing participant burden by limiting item redundancy. First, we identified a broad pool of existing validated surveys and scales relevant to mental health assessment (*Figure 1A*). Concurrently, we compiled a separate and independent inventory of mental health “constructs”, encompassing symptoms and other phenomena associated with depression and anxiety (*Figure 1B*). This inventory drew upon prior work in three established frameworks: the Hierarchical Taxonomy of Psychopathology (HiTOP)^18^, the National Institute of Mental Health’s Research Domain Criteria (RDoC)^19^, and the DSM-5 ^1^. We prioritized a list of specific constructs for this assessment protocol based on our consensus regarding their relevance to the aims and objectives of the study.

**Figure 1.**
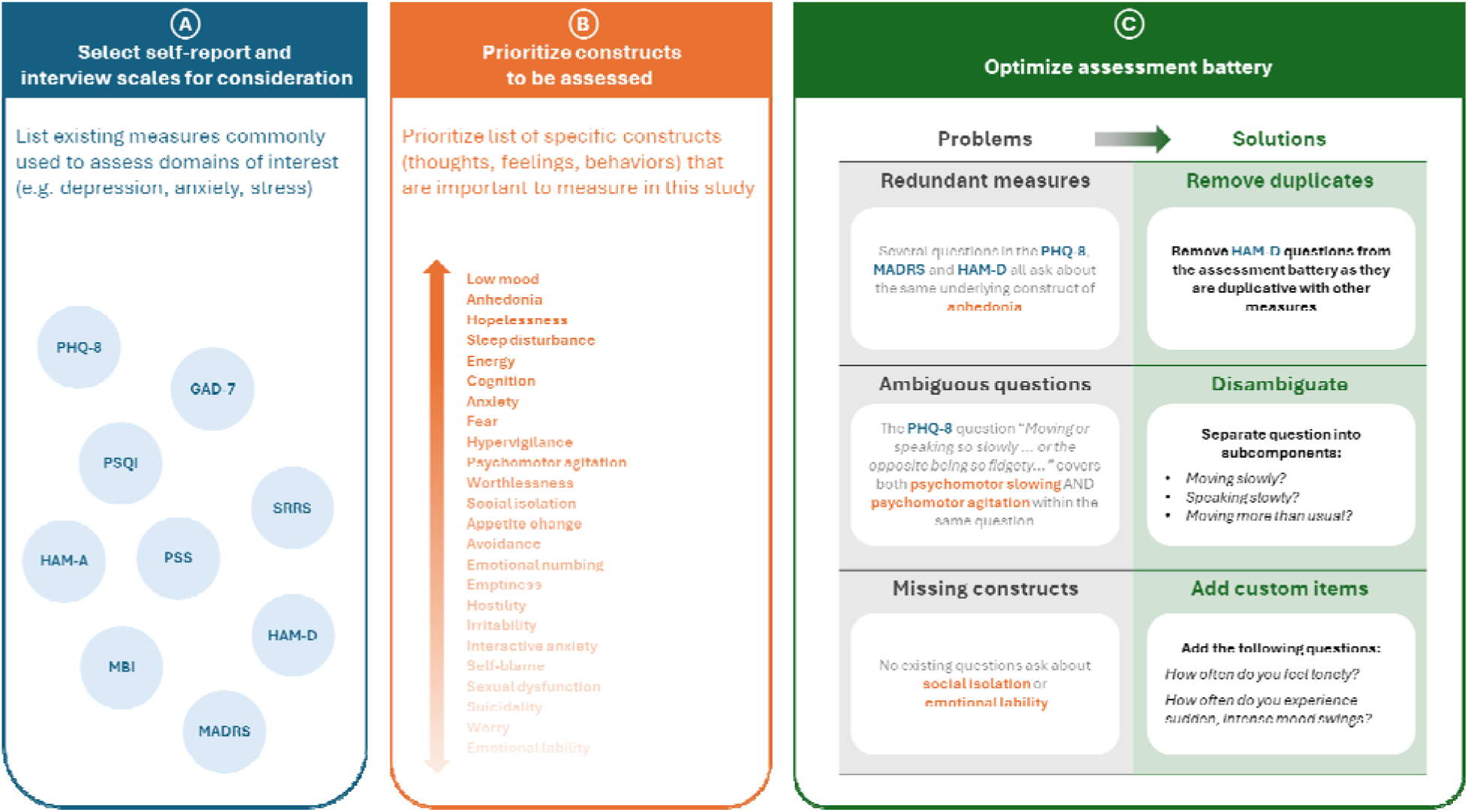
Schematic diagram of the three-step process used for developing the Digital Mental Health Study (DMHS) main study assessment protocol. **A.** We first selected for consideration a set of widely used validated interviews and self-report scales (key to abbreviations and citations below). **B.** We developed a separate prioritized inventory of mental health constructs (signs, symptoms, or other phenomena related to mental health), drawing on recent field-wide classification systems (HiTOP, RDoC and DSM-5). **C.** Each question in the potential instruments from Step A was then mapped to the construct from Step B which it covered. At this point, we could determine which constructs were receiving redundant assessment (i.e., assessed by multiple questions across different potential instruments) and which represented a gap in coverage (i.e., not assessed by any question across these instruments). We optimized the assessment protocol by minimizing redundant coverage, increasing the specificity of questions with ambiguous construct mappings, and adding sets of custom assessments to fill gaps in coverage. For example, the HAM-D showed similar overall construct coverage to the MADRS but with less specificity in each item, and therefore HAM-D was dropped, with the MADRS becoming the primary interview-based depression assessment. **Abbreviations:** CAT-MH: Computerized Adaptive Testing for Mental Health; GAD-7: Generalized Anxiety Disorder 7-item scale; MADRS: Montgomery-Åsberg Depression Rating Scale; SCID: Structured Clinical Interview for DSM Disorders; HAM-D: Hamilton Depression Rating Scale; HAM-A: Hamilton Anxiety Rating Scale; HiTOP: Hierarchical Taxonomy of Psychopathology; RDoC: Research Domain Criteria; DSM: Diagnostic and Statistical Manual of Mental Disorders.

Team members with expertise in depression and anxiety research then mapped individual questions from the above pool of survey instruments to the list of prioritized constructs. For example, the eighth item of the PHQ-8 (a query of “moving or speaking so slowly that other people could have noticed, or the opposite – being so fidgety or restless that you have been moving around a lot more than usual”) mapped to three distinct constructs: “psychomotor slowing: movement”, “psychomotor slowing: speech”, and “psychomotor agitation”. This comprehensive mapping allowed us to evaluate construct coverage across multiple potential instruments and to address three key issues. First, it revealed redundancies, suggesting assessments or items that could be removed to reduce burden without compromising essential information. Second, it identified survey questions that required disambiguation to achieve the specificity necessary for achieving study goals. Finally, it identified gaps in coverage of constructs that we deemed important to study goals but that were not assessed by existing instruments.

We leveraged this information in a final optimization step, employing a three-pronged approach (*Figure 1C*). First, we eliminated redundant instruments, prioritizing those with the clearest and most specific questions when constructs were covered by multiple instruments. Second, we supplemented existing questions with custom follow-up questions to disambiguate construct mappings. For example, we supplemented our version of the PHQ-8 by adding contingent questions to differentiate psychomotor agitation from psychomotor slowing of movement and psychomotor slowing of speech. Third, we added purpose-designed questions to address identified gaps (see *Monthly Symptoms and Events* section below), ensuring that all essential constructs were assessed (descriptions of all survey instruments are in *Supplementary Table 2*).

In establishing the final protocol (*Figure 2*), we selected the cadence for each survey based on expected symptom variability. For example, subjective experience of stress and current mood, given their potential for intra-day variability, were evaluated up to multiple times daily via ecological momentary assessment (EMA). We evaluated self-report symptoms of depression and anxiety (assessed by PHQ-8 and Generalized Anxiety Disorder 7-item scale [GAD-7]^20^) biweekly throughout the 52-week study. Variables that we expected to be relatively stable across the course of a year were measured less frequently (e.g., occurrence of stressful life events was assessed quarterly). The final study protocol encompassed self-report questionnaires, interviewer-administered rating scales, and collection of hair samples for potential measures of long-term hormonal stress response (details of study assessments are provided below). The estimated total time commitment for these assessments was approximately 37 hours over the 52 weeks. Each of these assessments is described in further detail below.

**Figure 2.**
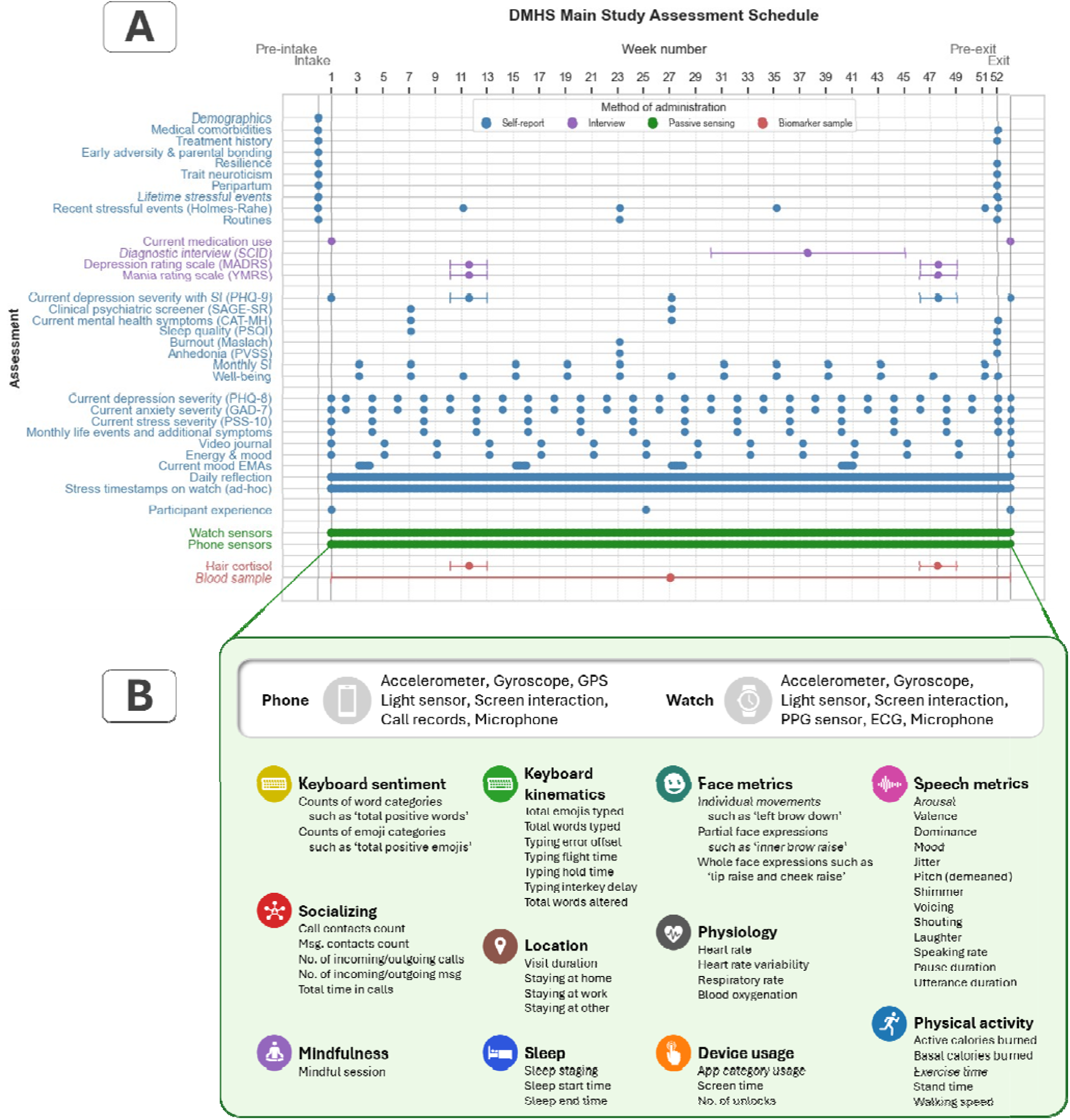
Overview of the final assessment protocol implemented in the DMHS. **A**. Depiction of the protocol, which spans a period of at least one year for every participant completing the study and encompasses a range of assessments including self-report measures, interviews conducted by trained staff, and continuous digital sensor streams (from iPhone and Apple Watch). **B.** Summary of key features collected in the passive data monitoring protocol, derived from the combined set of Apple Watch and iPhone data sensors. These features leveraged the Apple SensorKit and HealthKit data collection platforms. **Abbreviations:** MADRS: Montgomery-Åsberg Depression Rating Scale; YMRS: Young Mania Rating Scale; SCID: Structured Clinical Interview for DSM Disorders; SAGE-SR = Screening Assessment for Guiding Evaluation-Self-Report; CAT-MH: Computerized Adaptive Testing for Mental Health; PHQ-9: Patient Health Questionnaire-9; PSQI: Pittsburgh Sleep Quality Index; Maslach Burnout: Maslach Burnout Inventory; PVSS: Positive Valence Systems Scale; Holmes-Rahe: Holmes-Rahe Life Stress Inventory; PHQ-8: Patient Health Questionnaire-8; GAD-7: Generalized Anxiety Disorder 7-item scale; PSS-10: Perceived Stress Scale-10.

#### Depression and Anxiety Symptoms

We assessed depression and anxiety symptoms biweekly using the PHQ-8 and the GAD-7, respectively. Both surveys were administered remotely via the Apple Research app (except when administered via REDCap at the exit assessment). The full PHQ-9 (which adds a question on suicidality to the PHQ-8) was collected quarterly via REDCap. To disambiguate responses relating to psychomotor, sleep, appetite, and anhedonia constructs, we added contingent questions to both the PHQ-8 and PHQ-9 such that, if participants endorsed one of these items, they were asked to further clarify which specific symptoms they were experiencing (see *Supplementary Table 3A*).

To obtain a more comprehensive assessment of depression symptomatology than that obtained from these self-report surveys, trained raters administered the Montgomery-Åsberg Depression Rating Scale (MADRS)^21^ at two timepoints (approximately weeks 10-13 and 46-49), always on the same day that a PHQ-9 was obtained. While both self-report and interviewer-administered rating scales represent standard, widely utilized methods of symptom assessment, results from these different modalities do not consistently demonstrate high correlations.^22^ Each instrument may preferentially capture distinct features of depression; self-report scales such as the PHQ-9 emphasize assessment of an individual’s subjective experience of symptoms and emotional state, whereas interviewer-administered rating scales like the MADRS may better capture observable behaviors and symptoms that could be difficult for participants to accurately self-report. We chose to obtain these scales around the end of the 1^st^ quarter and middle of the 4^th^ quarter (rather than at intake and exit) to ensure that we could assess associations of these scales to sensor data collected both before and after the assessments.

#### Suicidality

A customized survey was administered that incorporated concepts from existing scales^23,24^ with the aim of distinguishing between passive and active suicidal ideation over the previous two weeks. This self-report assessment was administered every four weeks throughout the study via REDCap. In addition to providing research data, this survey identified participants with active suicidal ideation who warranted a follow-up assessment (see Participant Safety and Ethical Considerations, above).

#### Perceived Stress

Perceived stress was captured every four weeks via the Apple Research app using the Perceived Stress Scale-10 (PSS-10), which measures subjective appraisal of whether one’s life circumstances and events exceed one’s adaptive capacity.^25^ In addition, participants were asked to rate whether they felt stressed, as part of the Daily Reflection survey, and could record a Stress Log via the Apple Watch at any point during the study (see Supplementary Methods for details).

#### Exposure to stressful events

Early life adversity was assessed once using an Early Life Adversity and Parental Bonding questionnaire at the beginning of the study. Exposure to major stressful life events, which are typically considered moderators or predictors of mental health states,^26,27^ was assessed using a Lifetime History of Stressful Life Events survey (adapted from the List of Threatening Experiences^28^, Stressful Life Events Screening Questionnaire^29^ and the Life Events Checklist for DSM-5^30^) at the beginning and end of the study via REDCap; this survey was supplemented on a quarterly basis by the Holmes-Rahe Life Stress Inventory.^31^

#### Monthly Symptoms and Events

To assess symptoms relevant to depression and anxiety not measured in the surveys described above, we developed a custom survey consisting of 13 questions (some of which had follow-up queries) which we asked participants to answer every four weeks throughout the study via the Apple Research app *(Supplementary Table 3B*). To develop this custom survey, we identified all concepts in the construct inventory (*Figure 1B*) that did not map to any of the surveys that we were considering using. Then, three experts on depression and anxiety (ZDC, MGC and NF) ranked these concepts based on their judgment of their relevance to the study aims; those that demonstrated the highest degree of consensus between the three raters were selected for inclusion in the study.

#### Broad-based Clinical Assessments

We assessed participants for a broad range of mental health conditions beyond depression and anxiety. The Computerized Adaptive Testing – Mental Health survey (CAT-MH)^32^ is a self-report instrument that uses an adaptive algorithm to provide a rapid screen for current symptoms of depression, anxiety, mania/hypomania, post-traumatic stress disorder, and substance use disorders. We obtained this survey from all participants at weeks 7 and 27, and at exit via REDCap. Diagnostic information for major mental health disorders was obtained from all participants (at weeks 7 and 27) through completion of a comprehensive self-report behavioral health diagnostic screening tool, the Screening Assessment for Guiding Evaluation: Self-Report (SAGE-SR).^33^ All individuals who endorsed symptoms suggestive of mania or hypomania on the SAGE-SR or CAT-MH were further assessed for these conditions by a trained rater using the Young Mania Rating Scale (YMRS) at approximately weeks 10-13 and 46-49.^34^

We invited 240 participants for a structured diagnostic interview to confirm diagnosis of a current major depressive episode (MDE), selected based on inconsistent screening scores on the PHQ-8 (at week 26), PHQ-9, CAT-MH, and SAGE (all at week 27). These interviews consisted of administration by trained raters of NetSCID, a computerized version of the Structured Clinical Interview for the DSM-5 (SCID-5)^35^.

#### Daily Mood and Ecological Momentary Assessments (EMAs)

EMAs, representing subjective ratings of emotional states by participants while in their natural environment, may more accurately represent those states than the responses to retrospective self-report scales. We obtained EMAs using two schedules. Participants completed a Daily Reflection EMA via the Apple Research app throughout their participation in the study, rating overall mood over the past 24 hours: sadness, anxiety, stress, calm, energy, happiness (*Supplementary Table 4A*). Additionally, we obtained High Frequency Current Mood EMA “bursts” on a quarterly basis; these bursts consisted of self-ratings of immediate mood three times daily for seven consecutive days (*Supplementary Table 4A*).

#### Participant Experience

Participants completed self-report surveys capturing their feedback about their experience in the study at the beginning, middle, and end of the study.

#### Audiovisual Journals

To further develop metrics correlating mood state with facial and speech features, participants provided monthly video recordings via the Apple Research app on their iPhones. These recordings captured responses to self-administered questions probing recent emotional state, including general mood, instances of anger, stress, and sadness, as well as recent positive events (*Supplementary Table 4B*).

#### Other Audiovisual (AV) Data

10-minute semi-structured conversations with the study staff were collected, with consent from both the participant and the study staff. These were collected twice, immediately prior to the MADRS, at approximately weeks 10-13 and 46-49. The conversational prompts asked by raters are listed in *Supplementary Table 4C*. In addition, all clinical interviews were AV recorded for safety and quality purposes, and participants could opt to have their recordings used for the study.

#### Hair Cortisol and Blood Samples

Participants were offered the opportunity to provide a blood sample for future genetic analyses at any time during the study for additional compensation. A subset of participants was offered the opportunity to provide hair samples at an in-person check-in session, at approximately weeks 10-13 and 46-49, for additional compensation. Hair samples were collected by study staff to assay cortisol levels.

### Passive Digital Assessments

The DMHS employed an extensive passive data collection protocol (*Supplementary Table 5*), leveraging the capabilities of Apple’s HealthKit and SensorKit platforms to unobtrusively assess participants’ physiology, behavior, and emotional expression in daily life. HealthKit and SensorKit are data collection frameworks that grant access to a broad range of data streams from both iPhone and Apple Watch, after the participant authorizes sharing the data with the study. These frameworks employ on-device processing for many data streams to protect privacy. For example, rather than transmitting the raw GPS coordinates that have been used in prior digital phenotyping research, SensorKit algorithms categorize a person’s location directly on the phone and only report movements between anonymized locations. The raw location of participants is never shared or transmitted from either device.

The study protocol made use of many of the sensors on these devices (*Figure 2B*). Sensor data included accelerometer, gyroscope, and ambient light sensor readings from both iPhone and Apple Watch. Additionally, iPhone data included microphone input sampling (processed on-device for extraction of speech features such as pitch), screen time, call and text messaging metadata (such as duration and counts, excluding content of the calls), and application usage metadata (such as total duration of categories of apps). The Apple Watch also provided microphone input (similarly processed on-device), screen time, photoplethysmography (PPG) sensor data, and opportunistic electrocardiogram (ECG) sensor data.

From these diverse sensor data, over 200 behavioral and physiologic features were extracted via HealthKit and SensorKit, summarized here (see *Supplementary Table 5* for details).

- Mobility and location features, derived from iPhone location data, included visit duration at various anonymized locations and the total amount of time spent at home and work.
- Sleep-related features, such as sleep staging (REM, light, deep), sleep start time, and sleep end time, were derived using the Apple Watch’s on-device sleep annotation algorithm.
- Social interaction features, reflecting social engagement and communication patterns, were extracted from iPhone phone call and message metadata (excluding content of these communications), including call and message contact counts, the number of incoming and outgoing calls and messages, and total call duration, but excluding content of these communications.
- Speech features were derived from iPhone and Apple Watch microphone data through on-device processing, including measures of vocal prosody (pitch, jitter, shimmer, voicing, and speaking rate) as well as estimates of vocal emotion (valence, arousal, dominance, and mood) and specific vocal events (laughter, shouting).
- Data from the iPhone’s front-facing camera were used, with on-device processing, to derive measurements of facial expressions (such as eyebrow raise) after device unlock and during usage of messaging apps.
- Keyboard features, capturing typing dynamics from iPhone keyboard interactions, encompassed keyboard usage (frequency and duration), total emojis typed, total words typed, typing error offset, typing flight time, typing hold time, typing inter-key delay, and words altered. Sentiment analysis of typed text on the iPhone, also performed on-device, yielded features such as the frequency of absolutist words, anger emojis, anger words, anxiety emojis, anxiety words, confused emojis, and death-related words.
- Apple Watch PPG and ECG data provided vital signs, including heart rate, heart rate variability (HRV), respiratory rate, and blood oxygenation (SpO2).
- Physical activity features, such as calories burned, exercise time, stand time, and walking speed, were obtained from sensors on both devices.
- Finally, device usage patterns, including usage by app categories (duration, frequency), overall screen time, and the number of unlocks, were obtained from screen interaction data from both the iPhone and Apple Watch. Any user-initiated “mindful sessions” were also recorded as mindful minutes.

Participant privacy and data security were critically important during the design of this protocol. All data collection and processing procedures strictly adhered to relevant regulations including IRB oversight. In addition to providing informed consent, participants reviewed and authorized each data type on-device before any data were collected from their phone. Key safeguards included on-device processing to minimize the transmission of raw data. Data were deidentified before storage and analysis, utilizing unique identifiers separated from personally identifiable information. Industry-standard encryption protocols were employed for data both in transit and at rest. Secure servers with restricted access were used for data storage, following established best practices for data security.

### Medical History and Biological Markers

Current medication use and history of medical conditions were assessed at intake and exit. Pregnancy and childbirth history were assessed for female participants. For UCLA Health System patients, electronic health records were accessed for health conditions, hospitalizations, medications, and treatment history for five years prior to and during the study.

### Functional Academic Measurements

For participants who were current UCLA students, we obtained academic performance data including entrance exam scores, area of study, grades, and course enrollment.

### Data Monitoring

Survey data collected through the Apple Research app were continuously exported from the app and integrated into the study REDCap to allow comprehensive data monitoring by the study team. The degree to which participants adhered to the study protocol for wearing the Apple Watch was measured using HealthKit data that participants authorized to be shared with the study. An hour was treated as compliant for watch wear if data existed in the StandHour field (either 0 or 1) for that hour. These data were uploaded to Apple, processed, and then shared with the UCLA study team. Data completeness was rigorously monitored throughout the study, by members of the study team. Participants were informed at intake that the study team would contact them via email or phone regarding missing data to troubleshoot or provide reminders. Missing data criteria varied by type (e.g., three consecutive missing Daily Reflection surveys triggered a flag) (*Supplementary Table 6)*.

### Participant Compensation

Participants were compensated on a tiered system at the end of each quarter based on data completion (Base: 50-55%; Silver: 67-75%; Gold: 83-89%). Specifically, participants could earn up to $1070 in total for completing all components of the study over the course of one year. This included up to $240 for completing assessment visits, up to $560 for completing the majority of frequent and recurring remote assessments (e.g., Daily Reflection Survey and watch wear), up to $160 for completing the majority of infrequent remote assessments (including the High Frequency Current Mood EMA “bursts” and Stress Logs made on the Apple Watch), and up to $110 in bonuses at the mid-point and end of the study. Additional compensation was provided for optional assessments, including hair samples, blood samples, and the diagnostic interview. Participants could keep the Apple Watch upon meeting a minimum threshold for provision of data and adherence to visits, over the entire study.

### Statistical Analysis of Protocol Adherence and Self-Reported Outcomes

Chi-squared tests of independence were used to examine the association between participant categories (i.e., sex at birth, age, race and ethnicity, depression severity at intake, change in depressive severity) and study completion status. One-way analysis of variance (ANOVA) was used to assess the effect of depressive status at intake on mean watch wear rates. Spearman correlations were used to estimate the associations between different self-reported outcomes.

## Results

### Recruitment and Enrollment of Participants

Between January 2022 and January 2023, 17,150 individuals were screened, resulting in 5,684 eligible individuals consenting to participate in the study. From this group, 4,542 completed the initial intake assessment (*Supplementary Figure 1*), of whom 3,333 (73.4% of those completing the initial assessment) maintained their participation for the full 12 months including the exit assessment (*Table 1*). Three participants requested deletion of their data after completing the study; as such, intake data are presented for 4,539 total participants and 3,332 completers. The sample that completed the intake assessment met our pre-set targets for representation across categories defined by severity of depression symptoms, age, sex at birth, and self-reported race and ethnicity. The proportion of the sample that participated for the full 12-month study exceeded 65% for all age and race and ethnicity subgroups. This completion rate showed significant associations with age (range from 65.3% for 18-25 years to 90.2% for >65 years) and race and ethnicity (ranging from 69.8% for White, Hispanic to 75.8% for White, non-Hispanic, Table 1). Student status did not predict dropout rate when controlling for age (*p* = 0.944); however, student status and age were highly collinear, as most of our youngest participants were students.

**Table 1.**
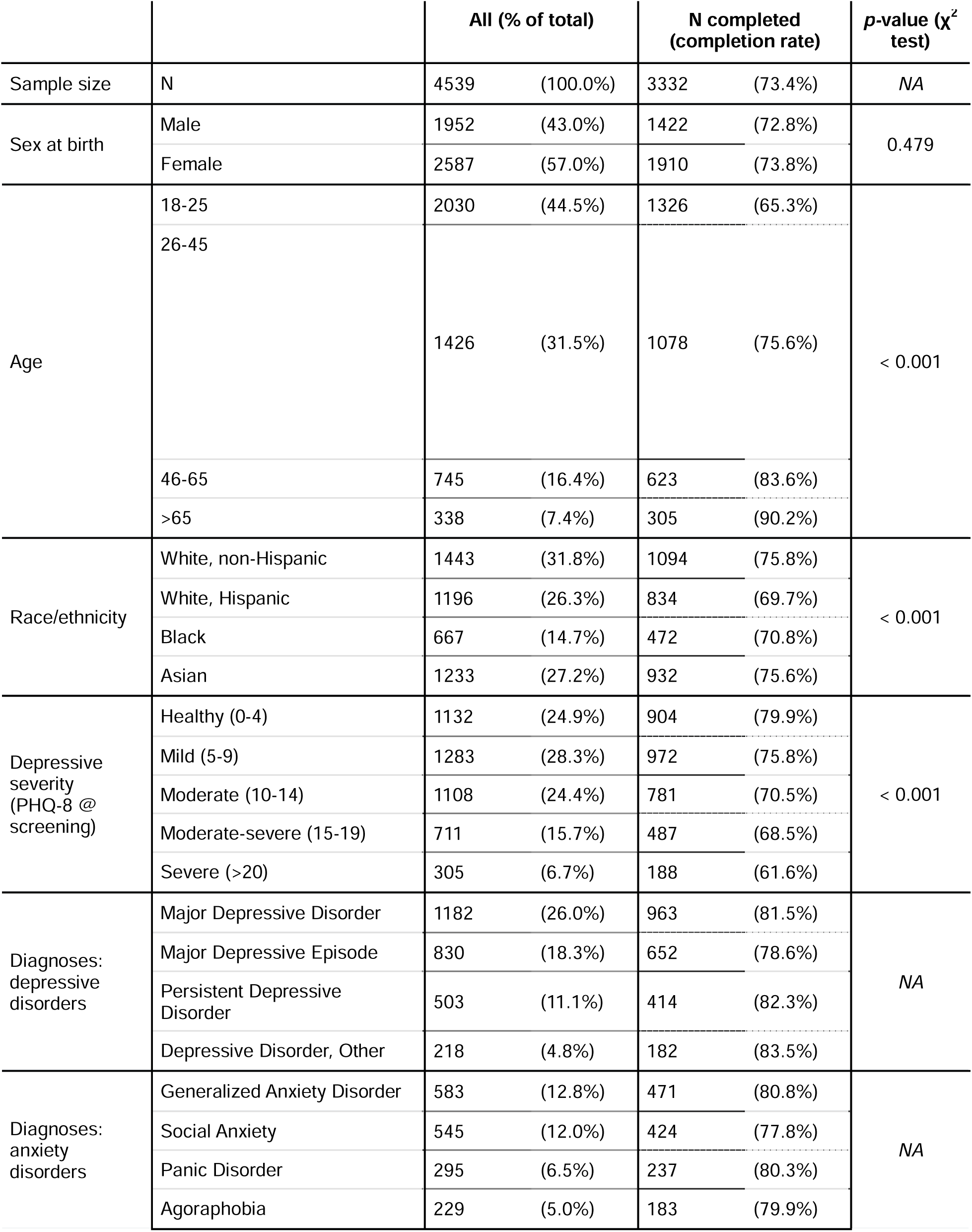
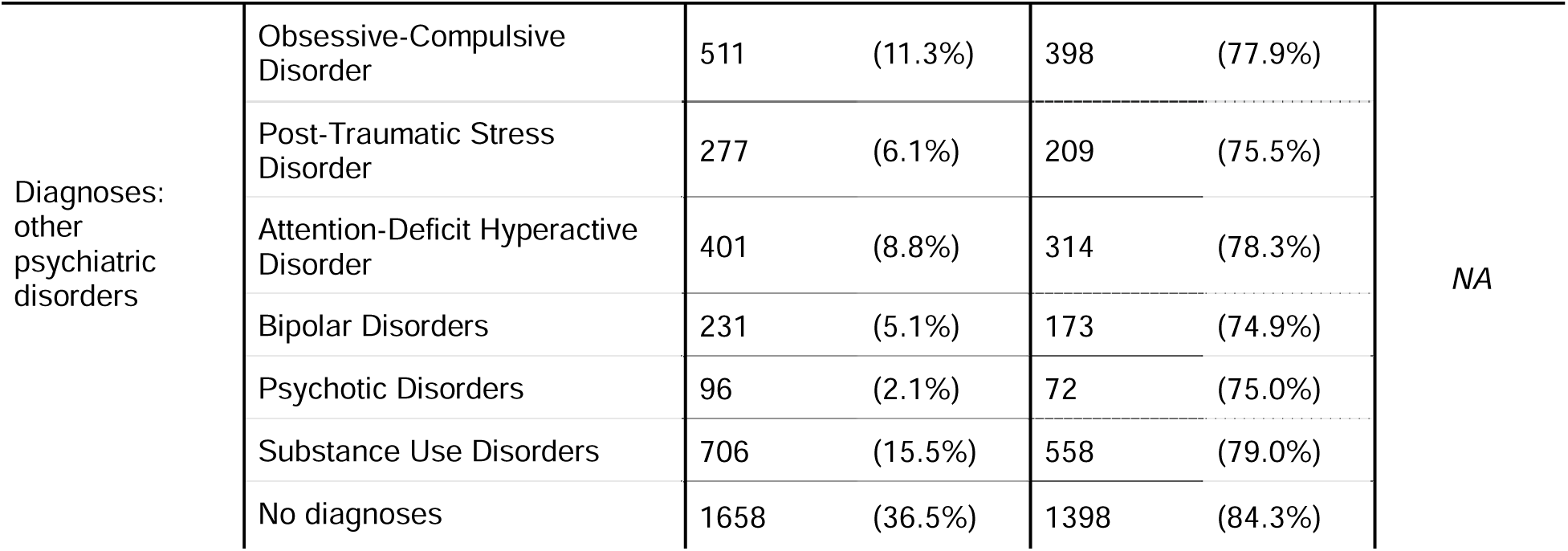
Participant characteristics in the DMHS. Recruitment bins were designed to achieve a diverse sample with respect to sex at birth, age, race/ethnicity, and depressive symptom severity at screening. Race and ethnicity were based on self-report at screening; participants identifying as Black regardless of Hispanic/Latino ethnicity were classified as Black, all other participants identifying as Hispanic/Latino ethnicity were classified as Hispanic/Latino, participants identifying as Asian and not Hispanic/Latino ethnicity were classified as Asian, and participants identifying as White, unknown or any other race and not Hispanic/Latino ethnicity were classified as Non-Hispanic White or Other. Depressive symptom severity at study entry was assessed using the PHQ-8. Diagnoses were derived using the SAGE-SR (at either of two timepoints when it was administered). To test whether subcategories differed from each other in study completion rate, separate Pearson’s Chi-squared tests for sex at birth, age, race and ethnicity, and depressive severity at screening were conducted (*p*-values indicated in last column). Abbreviations: PHQ-8: Patient Health Questionnaire-8; SAGE-SR = Screening Assessment for Guiding Evaluation-Self-Report.

The sample was selected to span the full range of depression symptom severity at intake, as determined by initial PHQ-8 score (*Table 1, Supplementary Figure 2*). Approximately half of the initial sample (47%, N=2,124) had a PHQ-8 score ≥10, a cutoff value associated with high sensitivity and specificity for capturing DSM-5 depression episodes. ^36^ Consistent with this expectation, the SAGE-SR suggested a likely depression diagnosis, at either of the two assessment time points, for 60.2% of participants. A substantial proportion of participants met criteria for likely anxiety disorders (36.4%) and other psychiatric disorders (48.9%), while 36.5% of participants did not qualify for any DSM-5 diagnosis according to SAGE-SR (*Table 1*). The completion rate exceeded 65% for all levels of intake depression severity and was significantly higher (*p* < 0.001) among those with an intake PHQ-8 < 10 (77.7%) compared to those with intake PHQ-8 >10 (68.5%).

### Longitudinal Trends in Study Dropout, Assessment Completion and Protocol Adherence

Overall assessment completion rates among those who finished the study were high, exceeding 85% for most tasks (*Figure 3A*, *Supplementary Table 7*), including the bi-weekly PHQ-8 (90.5%). Increases in dropout rates appeared to cluster around participation milestones, for example around Month 4, coinciding with the first check-in (weeks 10-13) and the end of the first compensation quarter *(Figure 3B*); this check-in was mandatory for continued participation. Dropout rates also increased around the time of the second check-in (weeks 46-49) and before the exit assessment, both of which were required for study completion.

**Figure 3.**
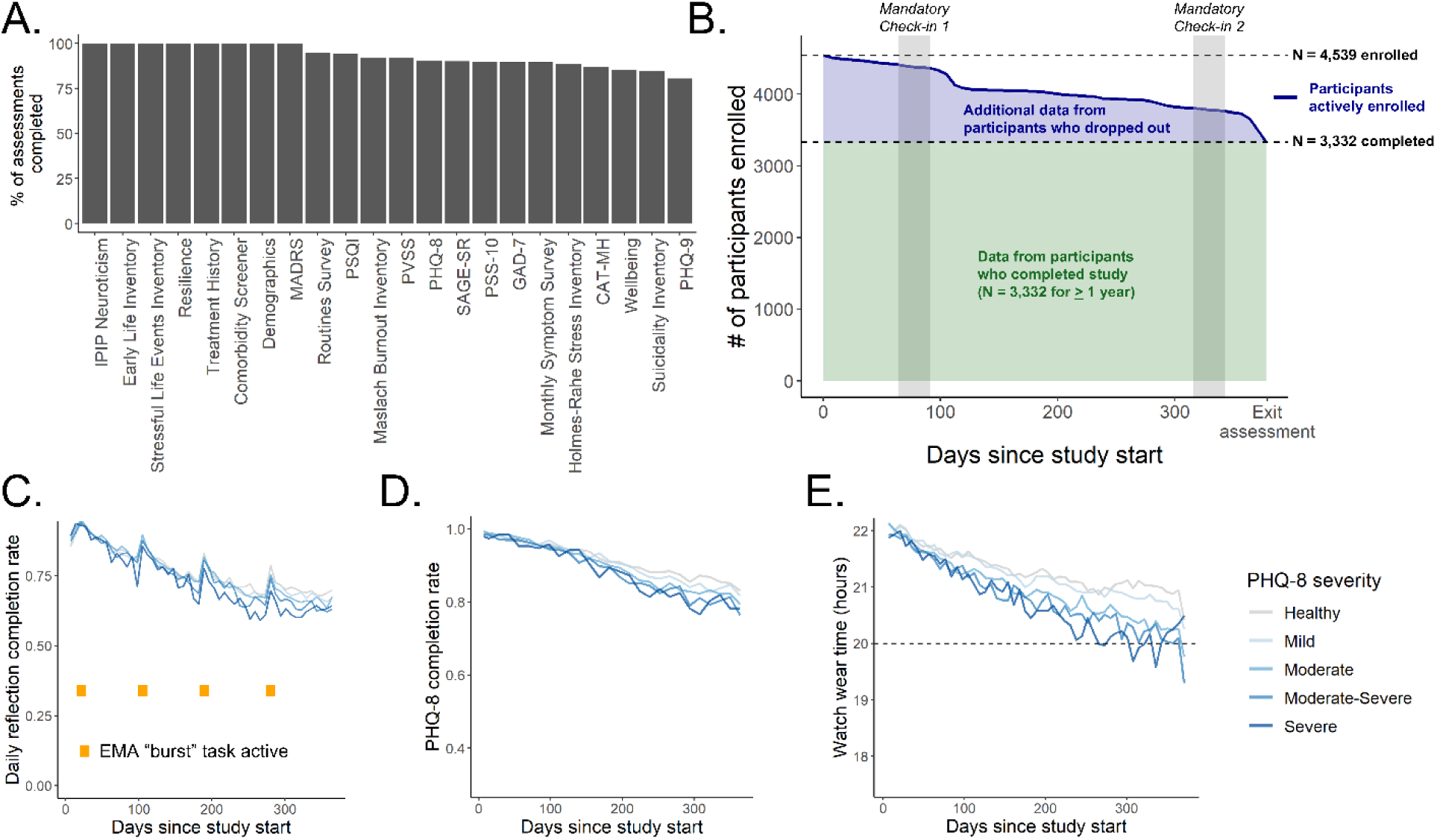
Overview of protocol adherence. **A.** Overall task adherence rates for individuals who participated for 12 months (“Completers”). **B.** Proportion of participants enrolled across the study duration, separated by depression severity (as measured by the PHQ-8) at screening. Highlighted bar indicates timing of mandatory initial check-in session, after which participants who could not finish the session were excluded from the remainder of the study. **C.** Adherence rate for the daily reflection survey task, separated by depression severity. Yellow bars indicate timing of ecological momentary assessment (EMA) “bursts”. **D.** Adherence rate for the biweekly PHQ-8 survey separated by depression severity. **E.** Mean watch wear time across the study duration, separated by depression severity. Dotted line indicates threshold for daily compliance (20 hours). **Abbreviations:** MADRS: Montgomery-Åsberg Depression Rating Scale (depression); PVSS: Positive Valence Systems Scale (hedonic functioning); PSQI: Pittsburgh Sleep Quality Index (sleep quality); IPIP: International Personality Item Pool (Neuroticism scale); SAGE-SR = Screening Assessment for Guiding Evaluation-Self-Report (psychiatric diagnosis); CAT-MH = Computerized Adaptive Testing-Mental Health (depression, anxiety); Patient Health Questionnaire-8 (depression); PSS-10: Perceived Stress Scale-10 (perceived stress); GAD-7: Generalized Anxiety Disorder 7-item scale (anxiety); PHQ-9: Patient Health Questionnaire-9 (depression).

Of the 1,207 participants who eventually dropped out, most remained enrolled past the halfway point of the study (median time to drop out of 241 days) (*Figure 3B*). For these individuals we continued to collect data from sensors, self-reports, and clinical interviews until the time that they formally dropped from the study roster.

Compliance with the Daily Reflection task (i.e., at least four daily reflection tasks completed per week) by study completers declined from a high of 86.9% in week 1 to 67.2% in week 52 (*Figure 3C*). Although baseline (BL) depression influenced the levels of such compliance (χ^2^(4) = 17.71, *p* = 0.001), these levels remained high throughout the participation period, even for the group with the most severe BL-depression symptoms (86.8% compliance, first 12 weeks; 72.8% overall) when compared to the no-BL-depression group (88.0% compliance, first 12 weeks; 77.4% overall). Daily completion rates were higher on weekdays (67.1%) than weekends (64.2%). Weeks with High Frequency Current Mood EMA bursts (see EMA burst, *Figure 3C*) showed a temporary boost in compliance rates, with a smaller carry-over effect.

Compliance with PHQ-8, among study completers (*Figure 3D*) was not significantly affected by BL depression severity (χ^2^(4) = 6.71, *p* = 0.152); while compliance rates among this group declined modestly throughout study, they remained above 80% through the final assessment. Mean watch wear time among completers (*Figure 3E*) declined slightly over the course of the study and was influenced by BL depression (F(4, 2485) = 4.98, *p* < 0.001); however, it remained above the 20-hour daily target for 87.4% of participants for the minimum number of days required for compensation (81.0% for severe BL depression, 88.4% for no BL depression).

### Longitudinal Symptom Trajectories

Repeated bi-weekly PHQ-8 assessments allowed for granular examination of depression symptom trajectories (*Supplementary Figure 3*). Overall, we observed initial improvement in scores during the first three months, followed by stabilization. This pattern was consistent across severity categories, with steeper improvement for higher BL severity. Such “early improvement” has been reported in other longitudinal studies.^37,38,39^

### Data Reliability: Correlational Analysis Across Standard Measures

To evaluate construct validity and provide an initial assessment of the reliability of survey responses in our study, we performed correlational analyses at both the aggregate scale level and the item level. Where possible, we compared these correlations against published results from prior population studies in the relevant domains.

At the scale level, we observed strong correlations between several of our core symptom assessment measures (*Figure 4A*). At intake, the correlation between total scores of PHQ-8 and GAD-7 was high (Spearman correlation = 0.753), consistent with prior research showing high comorbidity across the domains of depression and anxiety.^40^ The correlation between PHQ-9 (self-report) and MADRS (interview-based) conducted on the same day was also high (Spearman correlation = 0.811), in line with prior estimates.^41,42^ Perceived stress was highly correlated with both measures of depression (Spearman correlation = 0.84) and anxiety (Spearman correlation = 0.87), consistent with the hypothesis that such a perception may be a surrogate indicator of disrupted mental health. As expected, we observed inverse correlations between depression, anxiety, and perceived stress scales as compared against scales that assess positive psychological constructs (e.g., wellbeing, resilience, positive valence) (Spearman correlations ranging from -0.38 to -0.72).

**Figure 4.**
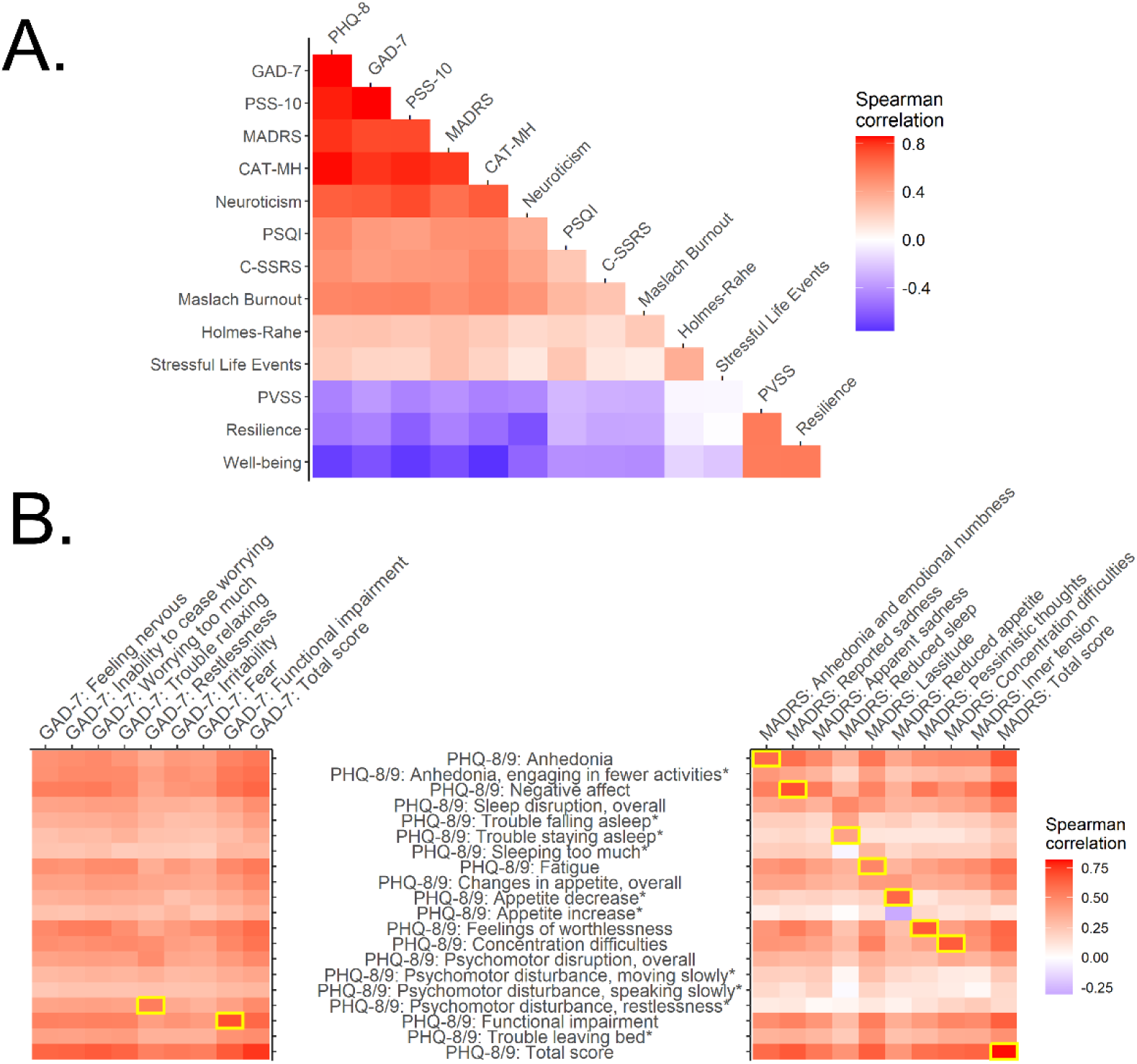
Relationships between scales in the DMHS. **A.** Spearman correlations between participant mean scores for all standard assessment scales administered. **B.** Spearman correlation between items of the PHQ-8 and GAD-7 at intake (left) and the first timepoint scores of the PHQ-9 and MADRS (right). Highlighted in yellow are pairs of items between the PHQ-8/9 and the GAD-7 and MADRS that assess overlapping symptom constructs. *indicates items that were added to the modified PHQ-8/9 scale utilized in DMHS. **Abbreviations:** PHQ-8/9: Patient Health Questionnaire-8/9; GAD-7: Generalized Anxiety Disorder 7-item scale; PSS-10: Perceived Stress Scale-10; CAT-MH: Computerized Adaptive Testing – Mental Health, Depressive Severity Score; Neuroticism: neuroticism sub-score of the International Personality Item Pool – Neuroticism, Extraversion, Openness; PSQI: Pittsburgh Sleep Quality Index; C-SSRS: Columbia-Suicide Severity Rating Scale; Maslach Burnout: Maslach Burnout Inventory; Holmes-Rahe: Holmes-Rahe Life Stress Inventory; PVSS: Positive Valence Systems Scale; Resilience: Connor-Davidson Resilience Scale.

To further assess construct validity in our core depression and anxiety instruments (MADRS, PHQ-8/PHQ-9, GAD-7), we conducted a more detailed item-level analysis across these instruments (see *Figure 4B*). We found item-level correlations to be substantially lower than correlations at the aggregate scale level, even when comparing the specific subset of items that are directly analogous between scales (e.g. *Figure 4B* highlighted in yellow). The recent OPTIMUM trial in geriatric depression analyzed item-level correlations between MADRS and PHQ-9 and thus provides a comparator for our study. The correlations between analogous items of the MADRS and PHQ-9 in our study (Spearman correlations ranging from 0.42-0.69, measured at weeks 10-13 of study) are very similar to those observed in the OPTIMUM trial (Spearman correlations ranging from 0.42-0.66 measured at week 10 of the study). ^43^ Additionally, both studies found the same four constructs of sadness, feelings of guilt/worthlessness, concentration difficulties, and suicidal thoughts to be most highly correlated across these two instruments, as compared to the remaining items.

Overall, the construct validity of our off-the shelf instruments are in line with those seen in other large population-based studies in the core domains of depression, anxiety and stress. This observation suggests that the reliability of participant responses in DMHS was comparable to that of other studies, despite the remote longitudinal data collection. The discrepancies between analogous MADRS and PHQ items seen both in our study and in literature, illustrate the importance of collecting both self-report and interviewer-administered ratings in our protocol.

### Contribution of Additional Purpose-Designed Assessments

The custom survey questions we added to our protocol to cover constructs missing from published instruments showed correlations in the moderate range with core depression, anxiety, and stress items (*Supplementary Figure 4*). This observation is consistent with our initial expectation that these questions would extract clinical information that is related (i.e. correlated) to core symptomology without being duplicative (i.e. without significant variance) compared to off-the-shelf items.

For items where the DMHS protocol added follow-up questions to disambiguate responses to the PHQ, we observed higher correlations with the analogous items in other scales. For example, the correlation between the MADRS item assessing appetite was higher with our disambiguated version of the analogous PHQ question (Spearman correlation = 0.61) versus the original ambiguous version in the PHQ (Spearman correlation = 0.41). Similarly, the GAD-7 item assessing restlessness had higher correlation with our disambiguated PHQ item (Spearman correlation = 0.54) versus the original ambiguous item on the PHQ (Spearman correlation = 0.48). These findings are consistent with the expectation that adding targeted follow-up questions to disambiguate these responses would improve construct validity of the symptoms being assessed.

### Participant Feedback

Feedback surveys at baseline (N=4,437), midpoint (week 25, N=3,388), and exit (N=3,332) indicated that most individuals considered study participation to be a generally positive experience (*Figure 5*). At baseline, 96% reported satisfaction with the intake interview, that study tasks were manageable, adequate compensation, confidence in compliance, and satisfaction with staff. Midpoint and exit surveys showed continued satisfaction, perceived benefit of mood logging, no major compliance issues, sufficient compensation, and manageable EMA burden. Visual inspection suggests that participants who dropped out, younger participants, and more depressed participants reported slightly greater difficulty adhering to tasks (*Supplementary Figure 5* - *Supplementary Figure 7*).

**Figure 5.**
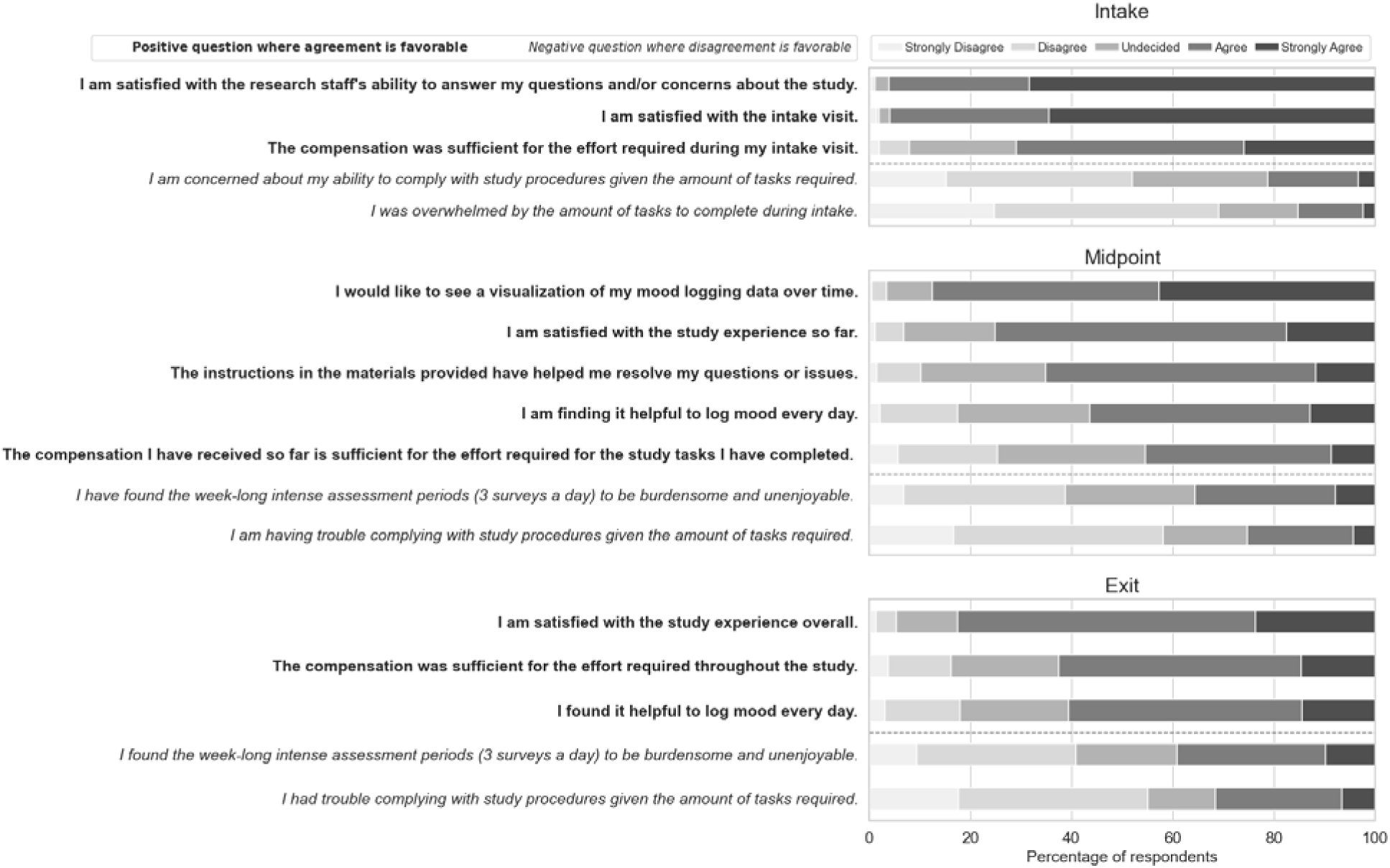
Overview of participant experience feedback at Intake, Midpoint (week 25), and Exit, provided by all participants who were enrolled and finished the survey at the time of administration. Items which are framed in the positive (i.e., agreement is favorable) are indicated with bold text while items framed in the negative (i.e., disagreement is favorable) are indicated with italics.

## Discussion

The DMHS aims to advance mental health research by demonstrating that data obtained passively from smartphones and smartwatches can provide phenotypes that objectively and precisely characterize physiological, behavioral, and emotional components of depression and anxiety, at a potentially enormous scale. This paper documents the project’s success in completing two foundational steps for achieving this objective. First, we demonstrated the feasibility of enrolling a sample of more than 4,000 individuals representing the full range of depression severity expected in the population and retaining most of them for a full year of intensive, and nearly entirely remote, data collection. Second, we established a protocol for obtaining a sufficiently broad range of digital phenotypes and symptomatic measures to capture the complexity and heterogeneity of depression and anxiety, and their relationship to both participants’ experience of stressful events and their subjective sense of their ability to cope with their life circumstances and events. The DMHS sample was more diverse (by age, self-described race, and ethnicity) and displayed a greater balance of males and females than previous mental health digital sensing studies.^44,45,46^ Most participants across these subgroups adhered to the assessment protocol throughout their participation in the study; we therefore anticipate that results from future DMHS analyses will be broadly generalizable.

Recruitment and retention of participants have been challenging for longitudinal studies of remote digital assessment in mental health, even those with a relatively limited burden of active assessment.^47^ The high proportion of participants who completed the full DMHS protocol (about 73% of the overall sample) is therefore noteworthy, given the intensive commitment it required (37 hours over the 12 months). Additionally, most participants who did not fulfill the requirements to be considered study completers, provided both sensor and survey data for nearly the entire study period before they dropped out. Therefore, these individuals will contribute to virtually all future analyses aimed at assessing associations between digital phenotypes and self-reported symptoms. As in previous digital mental health studies, retention rates were lower among younger participants and members of ethnic minority populations;^48,49^ however, these decrements were relatively small. Several possible factors could explain the relatively high retention rate in the DMHS compared to other studies: our employment of devices considered by most participants to be user-friendly (iPhones, which most already owned, and Apple Watches); proactive data monitoring and regular communication between study staff and participants; and manageable study burden and adequate compensation. The above information, as reported through direct participant feedback, suggests that relatively straightforward approaches can be used to sustain participant engagement even with extended and intensive assessment protocols. However, more systematic exploration of these and other potential factors influencing acceptance of longitudinal digital sensing will be necessary for this form of assessment to achieve integration in mental health care. From this standpoint, the high retention rate seen in DMHS participants who fulfill lifetime criteria for severe mental health diagnoses (including bipolar disorder and psychotic disorders) is highly encouraging.

The DMHS differs from prior digital phenotyping studies of mental health in its size and duration; the frequency and extensiveness of symptom assessments (by self-report scales, EMAs, and interviews); the wide range of digital sensors assayed, including sensor types (e.g., those assessing facial expressions), which have not previously been employed in this field; and the inclusion of multiple self-report and interview-based instruments plus purpose-designed questions not asked in standard surveys. We hypothesized that our strategy of selecting measures by systematically mapping survey items onto a unified inventory of mental health constructs could enable coverage of relevant domains while minimizing redundancies. The analyses that we undertook to evaluate correlations between different surveys suggest that we succeeded in achieving this initial objective; instrument level correlations were high, but the lower level of correlation between individual items (e.g., between those in self-report and interviewer-based depression scales) indicate the utility of the DMHS having employed such an extensive battery to test for associations to digital sensing features. This assessment battery required a substantial time commitment from participants that would likely not be feasible in efforts to further scale up mental health digital phenotyping studies. We therefore anticipate that a key aspect of DMHS analyses will be determination of the degree to which digital sensing data predict scores across different instruments; the results of these analyses may suggest the optimal approach for streamlining batteries for future studies.

Surprisingly few studies have conducted longitudinal evaluations of variability in depression and anxiety symptoms outside of mental health treatment settings; studies that have examined this topic have mostly been of short duration and with infrequent assessments.^50^ The 12-month duration of the DMHS offered the opportunity to detect clinically meaningful changes in symptomatology in a substantial proportion of participants. It is possible to time such major shifts in clinical state with a high degree of precision as we obtained PHQ-8 and GAD-7 assessments bi-weekly throughout the study; analyses of the temporal relation between digital sensing features and major shifts in clinical state can thus be conducted in the DMHS with a high degree of precision. Previous studies suggest, for example, that one possible mechanism underlying our observation of initial improvement followed by stabilization of depression scores could be that individuals have a greater propensity to enroll in research studies during periods when symptoms are more severe, and that after enrollment the distribution of symptom severity follows a regression to the mean.^51,52^

Among those participants who completed the full 12-month study, most continued to display a high degree of adherence to study tasks throughout its duration. The high completion rates across active and passive assessment modalities, in individuals across a broad range of diagnoses and symptom severities, suggests that patients may be able and willing to adhere to care plans that involve extended watchwear and data sharing. The decrease in adherence rate of daily mood EMA over the course of the study was greater than for other data types (although nearly half of participants continued to be adherent with these assessments on day 364), suggesting the utility of exploring alternative protocols for collecting this type of data. In designing such protocols, it is important to note that most participants reported that they considered it useful and non-burdensome to provide mood logging data.

Periodic assessment of participant attitudes towards digital assessments was an important component of the DMHS design. Most individuals reported, consistently, that participation was a positive experience, suggesting that research studies using extensive digital phenotyping may be broadly acceptable to potential participants.

### Limitations

We have identified four potential limitations of the DMHS. First, while the study sample was highly diverse with respect to multiple demographic and clinical variables, it did not attempt to achieve diversity in socioeconomic measures that may be important for both understanding depression and anxiety trajectories and usage patterns of digital devices; our use of approaches that depended on the internet may have limited our recruitment of individuals most affected by the socioeconomic “digital divide” in device usage.^53^ Additionally, all participants were recruited from the populations served by a single healthcare system and university, in a location (coastal Southern California) with minimal seasonal climate variation. This fact potentially limits the generalizability of findings related to behaviors (e.g., physical activity) that could be strongly affected by such seasonal variation. It is therefore essential that future studies attempt to replicate findings from DMHS in more diverse locations. Second, study participation was additionally limited to Apple iPhone users (or those willing to use a loaner iPhone) who may differ in unknown ways from individuals who use other devices. Third, we do not yet have an adequate understanding of the factors responsible for participants dropping out of the study, and therefore additional information is needed to design effective strategies to assess and then mitigate these factors. Finally, while we have detailed data about changes in symptomatology across the duration of the study, we did not systematically attempt to identify factors outside of the study that could contribute to such variation.

In summary, the DMHS represents a unique dataset, leveraging widely available and user-friendly consumer devices to demonstrate the feasibility of large-scale digital phenotyping and advance our understanding of common mental health conditions. Although analyses of this dataset are ongoing, we anticipate that they will demonstrate novel associations between a wide array of digital phenotype domains (including detailed assays of movement, sleep, social interaction, speech features, facial expressions, keyboard dynamics, and sentiment) and a set of symptomatic assessments that were more extensive and more frequently obtained than those in previous digital mental health studies. These association results may themselves contribute to more precise and objective frameworks for characterizing mental health symptoms and for unraveling the complexity and heterogeneity of mental health disorders. The results will also encourage further studies that will use phenotypes from digital devices to achieve even more ambitious aims. For example, the heterogeneity of depression and anxiety has presented an obstacle to efforts to identify specific genetic variants contributing to these conditions.^54,55,56^ Our experience with the DMHS indicates that it should be feasible to remotely obtain precisely defined and objective digital phenotypes for the much larger samples that would be needed for such genetic discovery studies. Similarly, we anticipate that that DMHS results will suggest immediate strategies for studies assessing the value of including digital phenotypes in clinical decision making and other precision treatment efforts.

## Declarations

## Acknowledgements

We thank the following individuals for their thoughtful discussion, helpful guidance, and other valuable efforts in support of this work: Jim Kretlow, Charles Wang, Athena Robinson, Arianna Yuan, Danielle Faruq, Lyndsie Slackie, Chris Godden, Michelle Popowitz, Antonia Petruse, and Doxa Chatzopoulou.

## Author Contributions

N.B.F., M.G.C., M.S.B., and J.F. conceived the project. N.B.F., M.G.C., and M.S.B. provided overall study leadership. They designed the protocol together with C.S.D., E.C., C.H., D.S., Z.D.C., and A.N. In addition, E.C. and C.H. were responsible for carrying out the protocol. R.A.B. provided protocol guidance throughout the study period. D.S., C.S.D., S.A., F.H., V.T., D.R-L., A.T.B., and B.B. contributed in conducting and interpreting analyses reported here. C.S.D., E.C., C.H., D.S., M.S.B., and N.B.F. constituted the primary writing group. All authors read and edited the manuscript.

## Data Availability

Data are not publicly available. Limited data access to support the findings of this paper may be available to research institutions. Any request for data access may be evaluated and responded to in a manner consistent with policies intended to protect participant confidentiality, language in the study protocol and informed consent form, and other applicable agreements with Apple. Requests for data access should be addressed to the corresponding author (N.B.F.).

