## Supplemental Materials for "Assessing the feasibility of large-scale digital sensing for depression and anxiety: The Digital Mental Health Study"

### SUPPLEMENTARY TEXT

**The DMHS Pilot Studies**

To assess the feasibility of plans for the DMHS we first performed two short pilot studies (Digital Mental Health Study Pilot 1 [DMHS-P1] and Digital Mental Health Study Pilot 2 [DMHS-P2]). We then applied lessons learned from these pilot studies in devising the DMHS protocol. The pilot studies had three main aims: (1) assess the feasibility of recruiting and retaining participants with different degrees of depression severity for longitudinal studies that passively collect digital sensing data from smartphones and smartwatches; (2) assess the feasibility of using recruitment protocols to achieve a study sample with pre-defined levels of diversity by age, sex at birth, and self-reported race and ethnicity; (3) evaluate the burden on participants of varying strategies for collecting extensive information from surveys and active tasks relating to symptoms of depression, anxiety, and stress symptoms. DMHS-P1 targeted recruitment of 140 individuals for an eight-week study while DMHS-P2 targeted recruitment of 120 individuals for a 12-week study. Both pilot studies recruited all their participants from among individuals registered as patients in UCLA Health and aimed to include participants evenly distributed by age (50% over 45), sex (50% female), self-reported ethnicity (25% each Asian, Black, Hispanic, and Non-Hispanic White or Other), and current depression severity, as measured with the self-reported Computerized Adaptive Testing for Mental Health (CAT-MH)[1] Depression severity score (50% experiencing none-to-mild depression symptoms and 50% experiencing moderate-to-severe depression symptoms).

**DMHS-P1**

In DMHS-P1, we used the CAT-MH to screen 1,744 prospective participants online between August and November 2020, most of whom either did not complete this screening or did not fulfill eligibility requirements (*Supplementary Figure 8*). Of those who were eligible and chose to enroll in the study (N=184), 164 participants completed at least the first assessment session, and 159 completed the final assessment session (*Supplementary Figure 8*). Eligibility criteria for DMHS-P1 are defined in *Supplementary Table 8*, the assessment schedule for DMHS-P1 is defined in *Supplementary Figure 9*, and study procedures are defined below.

Participants were randomly assigned to one of two study conditions with respect to Ecological Momentary Assessment (EMA) questions; one condition (which included 51% of the final sample) required completion of 10 daily questions while the other condition (49% of the final sample) required completion of only four daily questions (*Supplementary Table 10)***.** All other aspects of the protocol were uniform across the sample (*Supplementary Figure 9*); however, participants had the option to provide hair and saliva samples for potential biomarker analyses. The total time commitment for participants in DMHS-P1 ranged between 5.9 hours and 6.9 hours over the course of the two-month study period, reflecting the above two variables.

For this study, participants were asked to provide data for the 8-week study period, after the initial assessment session, using the provided Apple Watch and Beddit sleep tracker, and by using the Apple Research app installed on their personal iPhone or via online surveys sent throughout the study.

Overall retention rate was 96%, with 86% compliance rate for daily survey and watch wear, jointly on the same day. There was no significant difference between cohorts with and without depression symptoms, and no significant difference between the low and high burden study conditions. 87% were satisfied with the overall study experience.

The DMHS-P1 demonstrated to us that our overall approach was an efficient way to recruit and assess participants from diverse sociodemographic groups and across a wide range of depression severities. The high retention and compliance rates gave confidence that depressed patients could complete the protocol. Additionally, we determined that adoption of a relatively low burden EMA protocol could be important in achieving a high level of participant retention for a yearlong project (as envisioned for the DMHS main study).

**DMHS-P2**

For DMHS-P2, we screened 782 prospective participants online between March and May 2021, of whom 166 were eligible and enrolled in the study, 130 completed at least the first assessment session, and 121 completed the final assessment session (Supplementary Figure *10*).

Eligibility criteria for DMHS-P2 are defined in *Supplementary Table 9*, the assessment schedule for DMHS-P2 is defined in *Supplementary Figure 11*, and study procedures are described below. In DMHS-P2, participants were asked to complete mood ratings 4 times per day on the watch (*Supplementary Table 11*), in addition to regular tasks (as often as every two weeks) administered through the Research app installed on their phone (*Supplementary Figure 11*). Notable differences between the DMHS-P1 and DMHS-P2 protocols included the addition of audiovisual journals (see *Methods: Study Assessments*) and a structured diagnostic interview (NetSCID) incorporated in the intake assessment session for all participants in the latter study. The addition of the diagnostic interview lengthened this intake session substantially, from approximately 3 hours in DMHS-P1 to approximately 5.5 hours in DMHS-P2. Furthermore, DMHS-P2 participants were issued two watches and asked to wear a watch continuously throughout the study duration (wearing one watch while the other charged, including overnight). The time commitment for participants enrolled in DMHS-P2 was approximately 12 hours over the study period (12-14 weeks).

Participants were asked to provide data for the 12-week study period, after the initial assessment session, using the provided study equipment, which included two Apple Watches and the Beddit sleep tracker. Participants were asked to always wear one of the two Apple Watches for continuous day and night data collection and to use the Beddit sleep tracker for the 12-week study period. Participants were again asked to complete tasks using the Apple Research app installed on their personal iPhone or via online surveys sent throughout the study. In addition to the longer study duration, new assessments were added to the study protocol following DMHS-P1. Participants completed weekly Video Journals via the Apple Research app installed on their phone; participants had up to five minutes to record themselves responding to the following three prompts: “1 How are you feeling today? 2 Did you feel angry, stressed, or sad this week? If so, why? 3 Did something good happen this week? If so, what was it?”. Participants were asked to provide ad hoc mood timestamps using the Apple Watch. In addition to daily symptom assessments (where all participants now received 4 questions daily, *Supplementary Table 11*), participants were asked to complete a weekly symptom survey via the Apple Research app. In addition to the clinical rating scales to assess depression and anxiety, all participants also completed a structured diagnostic interview (SCID) as part of their intake assessment session at the beginning of the study. In this phase, half of the sample was selected to be offered the option of providing hair or saliva samples near the intake and exit assessment sessions for the measurement of cortisol for additional compensation. Sample collection kits were shipped to participants and returned to the research team by mail.

Based on DMHS-P2 we determined that an interviewer-based diagnostic assessment (i.e., NetSCID) was not scalable for the full DMHS and therefore decided to use a self-report DSM-5 diagnostic assessment (SAGE-SR) and reserve the NetSCID for a selected sub-sample (as described in *Methods*). Additionally, we decided that a watch-wearing protocol based on the use of two Apple Watches was too complex, and therefore instituted the simplified protocol described in *Methods*. Participants reported that they preferred to have all their tasks on one device, with one place to track completion. The EMA task was therefore reverted to the iPhone for the main DMHS study, with an optional watch survey task for bonus compensation (Stress Logs). We also learned that the Video Journal prompts often elicited very short responses (with means 15, 27, and 24 seconds). Questions 1 and 3 were therefore updated to the wording shown in *Supplementary Table 4B* to elicit longer responses in the main DMHS study.

### SUPPLEMENTARY FIGURES

### Supplementary Figure 1

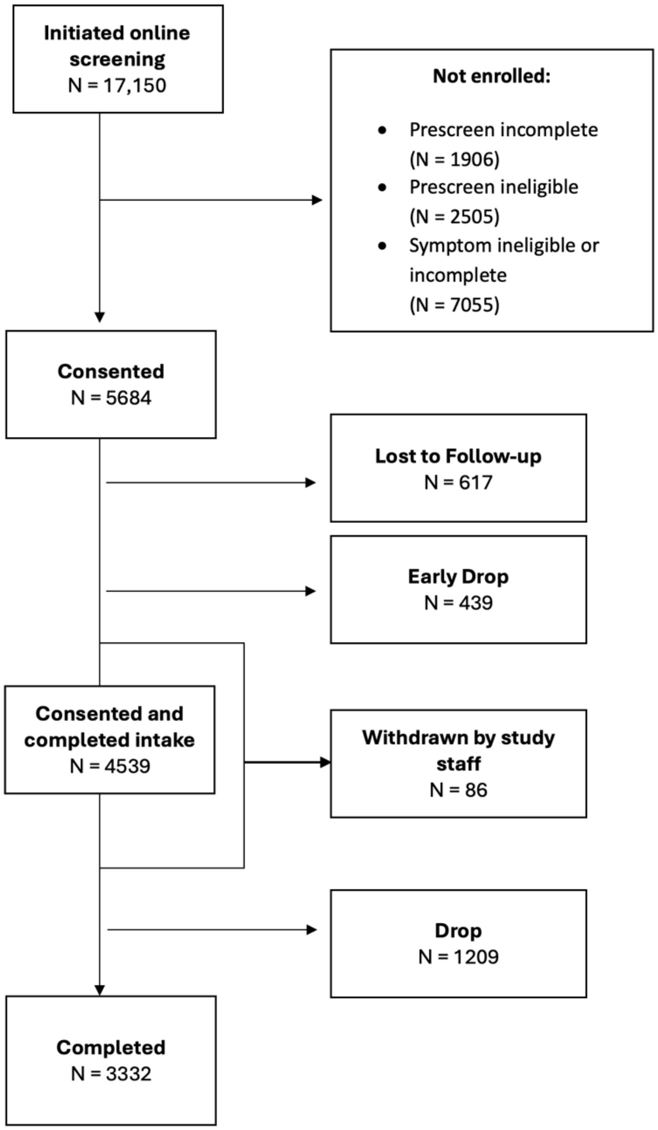


***Supplementary Figure 1*** Enrollment flowchart for the Digital Mental Health Study (DMHS). Lost to Follow-up: Non-responsive to study staff after enrollment, no assessments completed. Early Drop: Communicated reason for dropping from study before intake session or dropped before the entire intake session completed. Withdrawn by study staff: Determined that participant did not meet eligibility requirements after enrollment or was removed from the study for reasons other than non-responsiveness or non-compliance. Drop: Communicated reason for dropping from the study after completing intake session or the participants became non-responsive for scheduling remaining assessments and/or providing data during remote collection period. Completed: Completed all assessment sessions.

#### SUPPLEMENTARY FIGURE 2

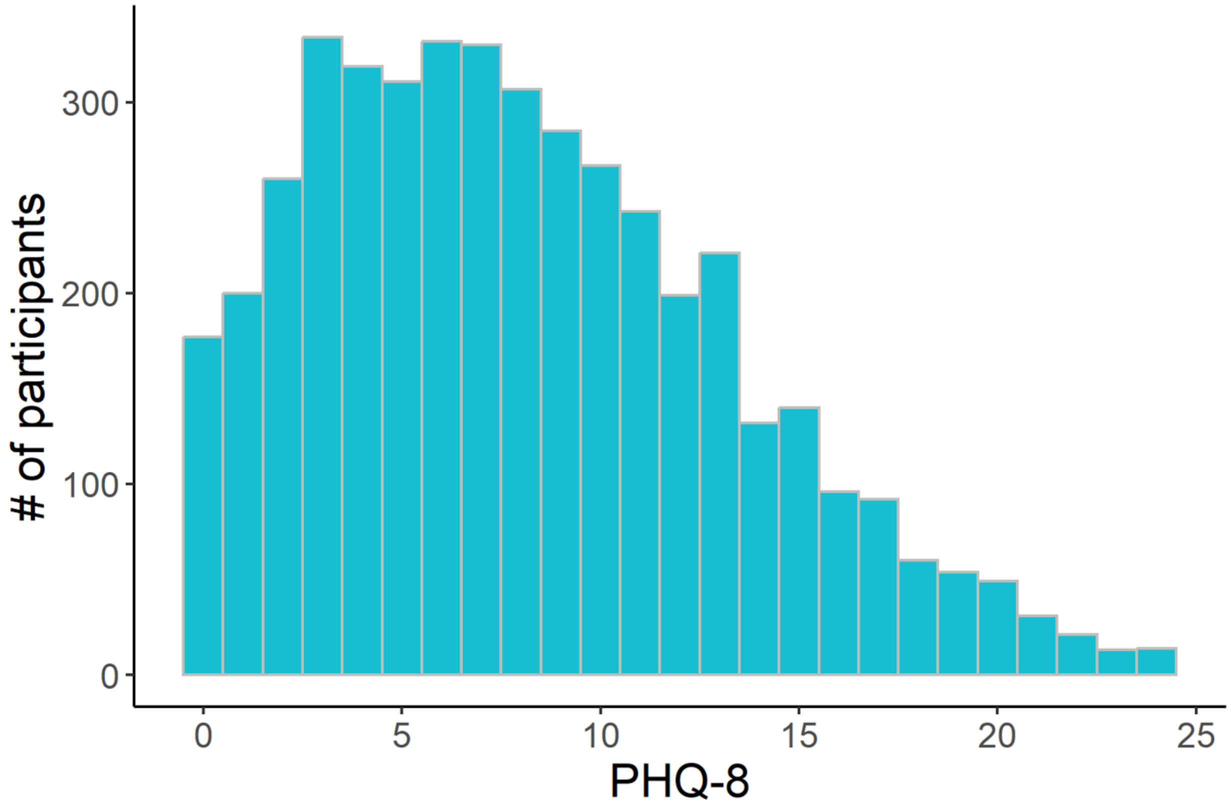


***Supplementary Figure 2*** Distribution of PHQ-8 scores at screening for all enrolled participants (N = 4539). **Abbreviations**: PHQ-8: Patient Health Questionnaire-8.

#### SUPPLEMENTARY FIGURE 3

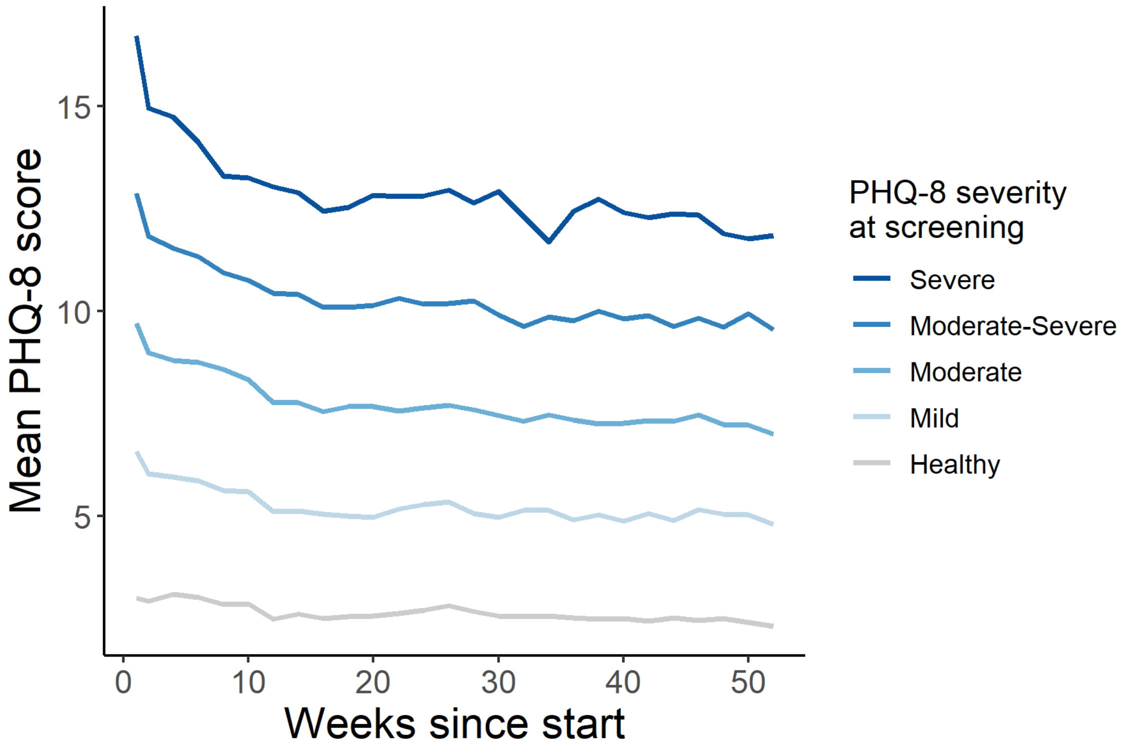


***Supplementary Figure 3*** Mean trajectories of PHQ-8 total score, separated by PHQ-8 severity at intake for all enrolled participants (N = 4539). **Abbreviations:** PHQ-8 = Patient Health Questionnaire-8.

#### SUPPLEMENTARY FIGURE 4

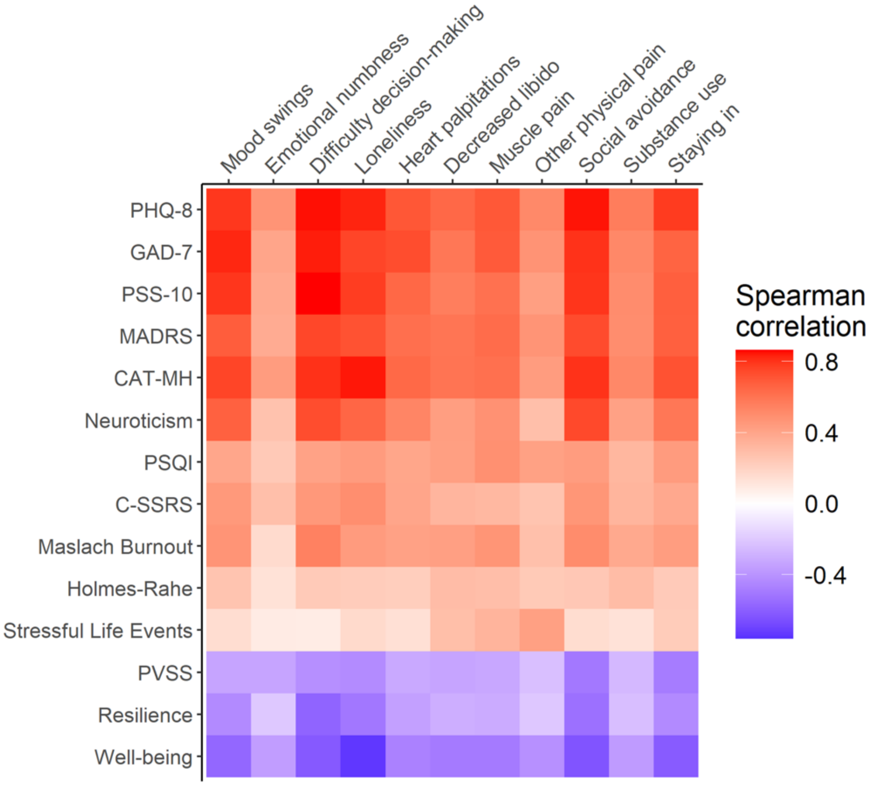


***Supplementary Figure 4*** Spearman correlations between all standard assessment scales (X-axis) and all custom assessment (Y-axis), which were selected to cover key gaps in construct coverage. **Abbreviations**: PHQ-8: Patient Health Questionnaire-8; GAD-7: Generalized Anxiety Disorder 7-item scale; PSS-10: Perceived Stress Scale-10; MADRS: Montgomery-Åsberg Depression Rating Scale; CAT-MH: Computerized Adaptive Testing for Mental Health Depressive Severity Score; Neuroticism: Neuroticism sub-score of the International Personality Item Pool – Neuroticism, Extraversion, Openness; PSQI: Pittsburgh Sleep Quality Index; C-SSRS: Columbia-Suicide Severity Rating Scale; Maslach Burnout: Maslach Burnout Inventory; Holmes-Rahe: Holmes-Rahe Life Stress Inventory; PVSS: Positive Valence Systems Scale; Resilience: Connor-Davidson Resilience Scale.

#### SUPPLEMENTARY FIGURE 5

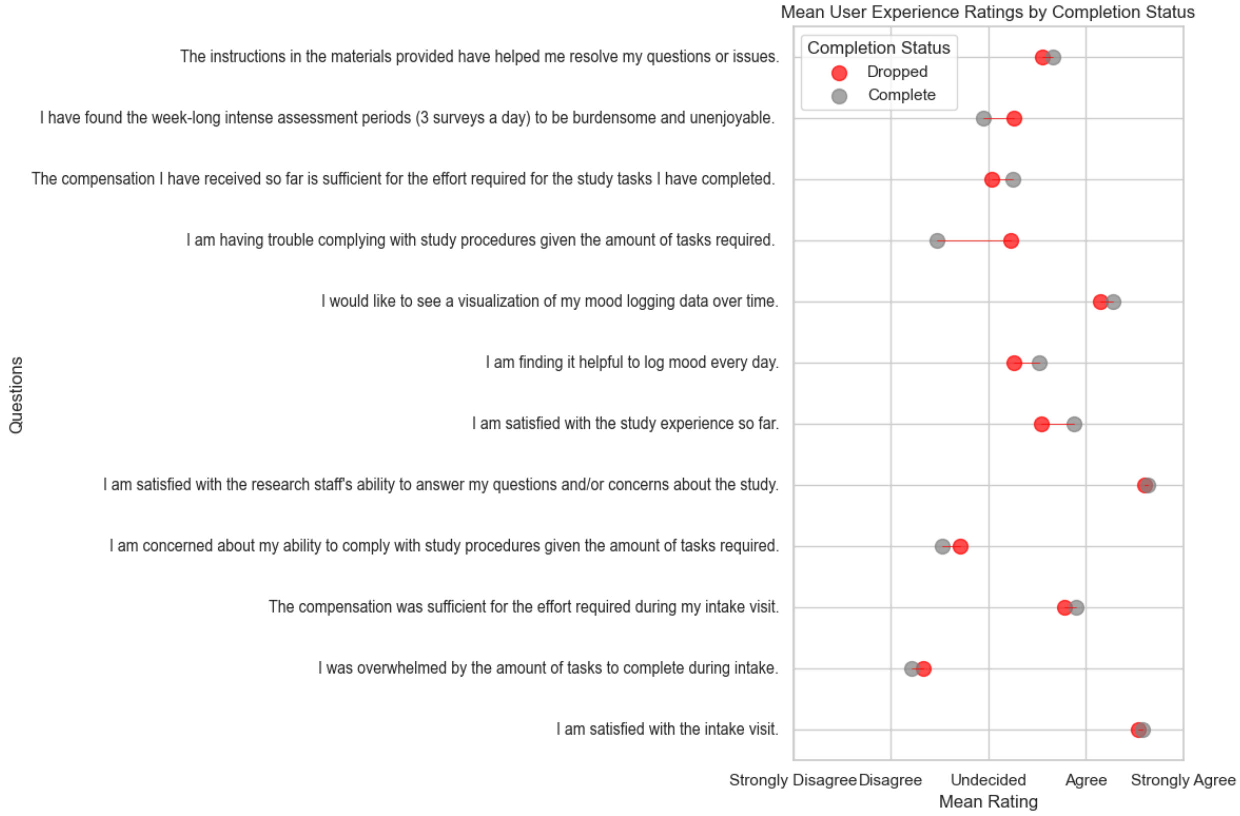


***Supplementary Figure 5*** Comparison of average user experience rating of those who successfully completed the DMHS study (grey) vs those who dropped out at some point prior to completion (red). Participants who dropped out showed higher average rating for difficulty complying with study procedures due to volume of tasks required.

#### SUPPLEMENTARY FIGURE 6

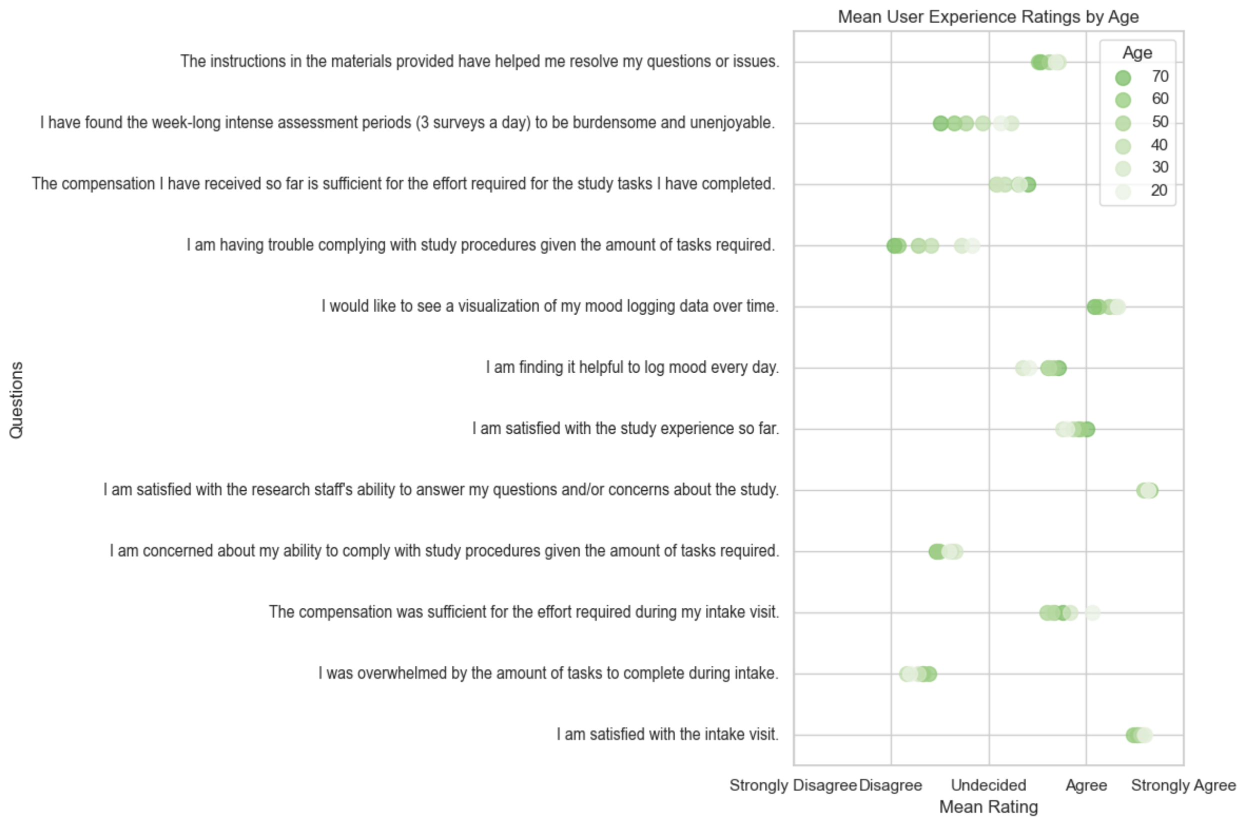


***Supplementary Figure 6*** Comparison of DMHS average user experience rating by age (nearest decade), younger in light green and older in dark green. Participants over 75 excluded from this chart due to low N. Older participants rated less burden from study assessments and less difficulty complying with study procedures**.**

### SUPPLEMENTARY FIGURE 7

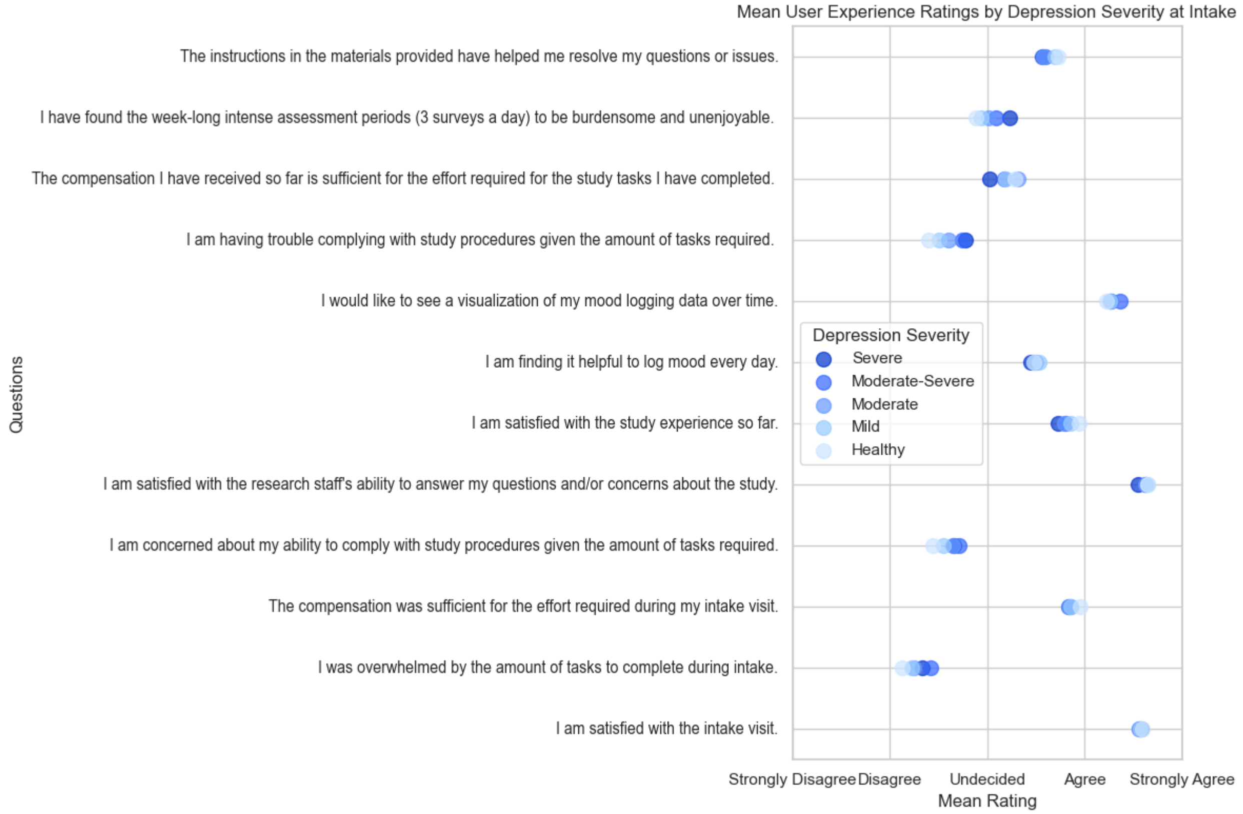


***Supplementary Figure 7*** Comparison of average user experience rating by depression severity at intake. Differences were relatively mild compared to those seen with final completion status and age, but those with more severe depression rated somewhat higher trouble completing study procedures.

#### SUPPLEMENTARY FIGURE 8

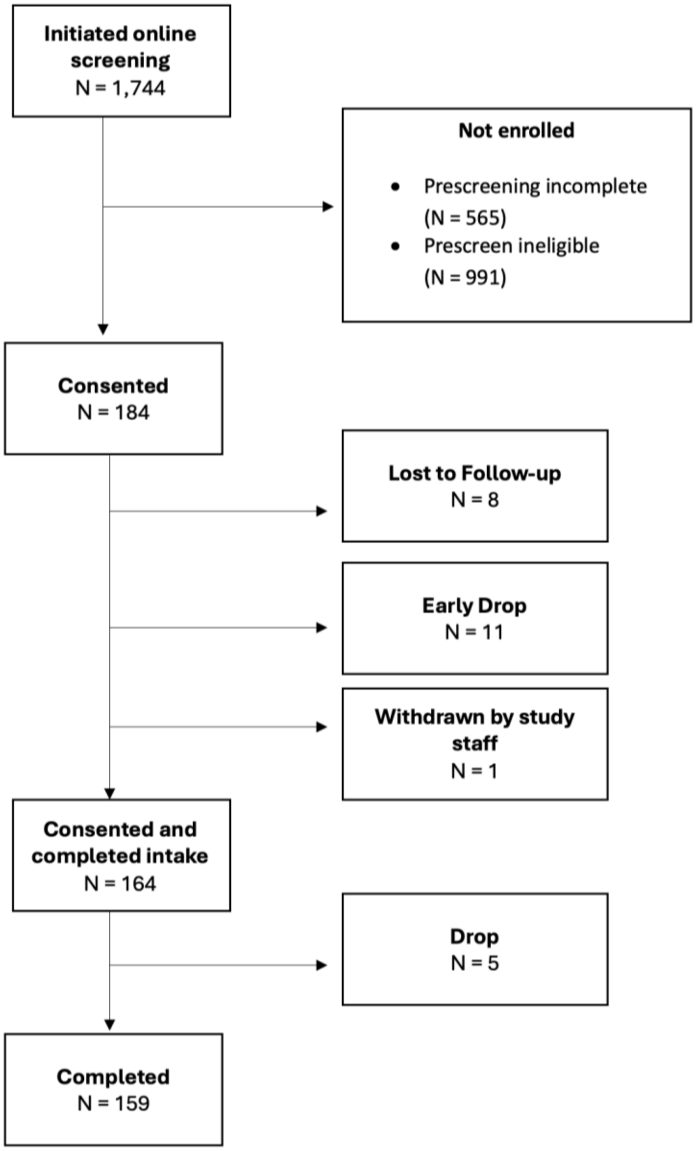


***Supplementary Figure 8*** Enrollment flowchart for the DMHS Pilot 1 (DMHS-P1) Study. Lost to Follow-up: Non-responsive to study staff after enrollment, no assessments completed. Early Drop: Communicated reason for dropping from study before intake session or dropped before the entire intake session completed. Withdrawn by study staff: Determined that participant did not meet eligibility requirements after enrollment or was removed from the study for reasons other than non-responsiveness or non-compliance. Drop: Communicated reason for dropping from the study after completing intake session or the participants became non-responsive for scheduling remaining assessments and/or providing data during remote collection period. Completed: Completed all assessment sessions.

### SUPPLEMENTARY FIGURE 9

| Task/Survey | Pre-Intake | Intake | W2 | W4 | W6 | W8 | Exit |
|---|---|---|---|---|---|---|---|
| Demographics | ✓ | | | | | | |
| Medical comorbidities | ✓ | | | | | | |
| Treatment history | ✓ | | | | | | ✓ |
| Early adversity & parental bonding | ✓ | | | | | | |
| Lifetime stressful events | ✓ | | | | | | ✓ |
| Recent stressful events (Holmes-Rahe) | ✓ | | | | | | ✓ |
| Big Five Personality Traits | ✓ | | | | | | |
| Resilience | ✓ | | | | | | |
| Routines | ✓ | | | | | | ✓ |
| Sleep quality (PSQI) | ✓ | | | | | | ✓ |
| Clinical psychiatric screener (SAGE-SR) | | ✓ | | | | | ✓ |
| Depression rating scale (MADRS) | | ✓ | | | | | ✓ |
| Anxiety rating scale (HAM-A) | | ✓ | | | | | ✓ |
| Current medication use | | ✓ | | | | | ✓ |
| Hair cortisol | | ✓ | | | | ✓ | |
| Salivary cortisol | | ✓ | | | | ✓ | |
| Current mental health symptoms (CAT-MH) | | ✓ | ✓ | ✓ | ✓ | ✓ | |
| Current depression severity (PHQ-8) | | ✓ | | ✓ | | ✓ | |
| Current anxiety severity (GAD-7) | | ✓ | | ✓ | | ✓ | |
| Current stress severity (PSS-10) | | ✓ | | ✓ | | ✓ | |
| Chronic stress ratings | | ✓ | | ✓ | | ✓ | |
| Disability scale (SDS-5) | | ✓ | | ✓ | | ✓ | |
| Participant experience | | ✓ | | ✓ | | | ✓ |
| Symptom survey | | ✓ | | | | ✓ | |
| Daily mood survey (4-item or 10-item) | | | | | | | |
| Watch sensors | | Daily | | | | | |
| Beddit sensors | | | | | | | |

***Supplementary Figure 9*** Assessment schedule for the DMHS-P1 Study. **Abbreviations:** Holmes-Rahe: Holmes-Rahe Life Stress Inventory; PSQI: Pittsburgh Sleep Quality Index; SAGE-SR = Screening Assessment for Guiding Evaluation-Self-Report; MADRS: Montgomery-Åsberg Depression Rating Scale; HAM-A: Hamilton Anxiety Rating Scale; CAT-MH: Computerized Adaptive Testing for Mental Health; PHQ-8: Patient Health Questionnaire-8; GAD-7: Generalized Anxiety Disorder 7-item scale; PSS-10: Perceived Stress Scale-10; SDS-5: Sheehan Disability Scale.

#### SUPPLEMENTARY FIGURE 10

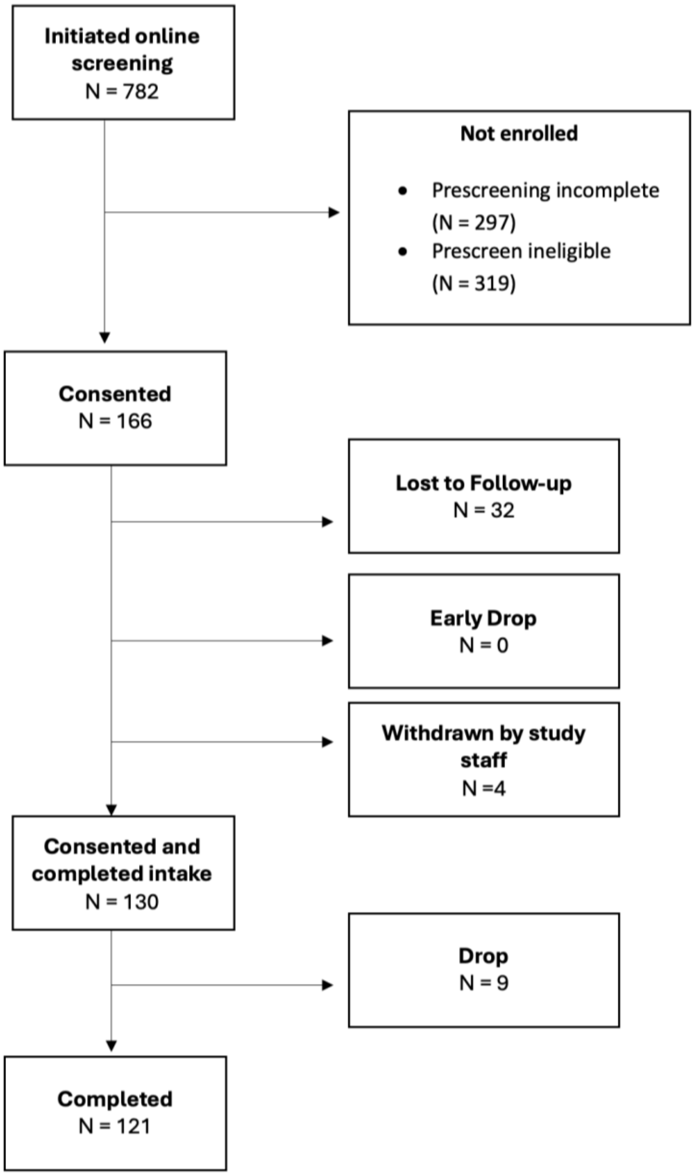


***Supplementary Figure 10*** Enrollment flowchart for the DMHS Pilot 2 (DMHS-P2) Study. Lost to Follow-up: Non-responsive to study staff after enrollment, no assessments completed. Early Drop: Communicated reason for dropping from study before intake session or dropped before the entire intake session completed. Withdrawn by study staff: Determined that participant did not meet eligibility requirements after enrollment or was removed from the study for reasons other than non-responsiveness or non-compliance. Drop: Communicated reason for dropping from the study after completing intake session or the participants became non-responsive for scheduling remaining assessments and/or providing data during remote collection period. Completed: Completed all assessment sessions.

#### SUPPLEMENTARY FIGURE 11

| Task/Survey | Pre-Intake | Intake | W1 | W2 | W3 | W4 | W5 | W6 | W7 | W8 | W9 | W10 | W11 | W12 | W13-W14[a] | Exit |
|---|---|---|---|---|---|---|---|---|---|---|---|---|---|---|---|---|
| Demographics | ✓ | | | | | | | | | | | | | | | |
| Medical comorbidities | ✓ | | | | | | | | | | | | | | | |
| Treatment history | ✓ | | | | | | | | | | | | | | | ✓ |
| Early adversity & parental bonding | ✓ | | | | | | | | | | | | | | | |
| Lifetime stressful events | ✓ | | | | | | | | | | | | | | | ✓ |
| Recent stressful events (Holmes-Rahe) | ✓ | | | | | | | | | | | | | | | ✓ |
| Trait neuroticism | ✓ | | | | | | | | | | | | | | | |
| Resilience | ✓ | | | | | | | | | | | | | | | |
| Routines | ✓ | | | | | | | | | | | | | | | ✓ |
| Sleep quality (PSQI) | ✓ | | | | | | | | | | | | | | | ✓ |
| Burnout (Maslach) | ✓ | | | | | | | | | | | | | | | ✓ |
| Peripartum | ✓ | | | | | | | | | | | | | | | ✓ |
| Life satisfaction | ✓ | | | | | | | | | | | | | | | ✓ |
| Clinical psychiatric screener (SAGE-SR | | ✓ | | | | | | | | | | | | | | ✓ |
| Depression rating scale (MADRS) | | ✓ | | | | | | | | | | | | | | ✓ |
| Anxiety rating scale (HAM-A) | | ✓ | | | | | | | | | | | | | | ✓ |
| Current medication use | | ✓ | | | | | | | | | | | | | | ✓ |
| Hair cortisol | | ✓ | | | | | | | | | | | | | | ✓ |
| Salivary cortisol | | ✓ | | | | | | | | | | | | | | ✓ |
| Diagnostic Interview (Net-SCID) | | ✓ | | | | | | | | | | | | | | |
| Current mental health symptoms (CAT-MH) | | ✓ | | ✓ | | ✓ | | ✓ | | ✓ | | ✓ | | | | ✓ |
| Current depression severity (PHQ-8) | | ✓ | | | ✓ | | ✓ | | ✓ | | ✓ | | ✓ | | | ✓ |
| Current anxiety severity (GAD-7) | | ✓ | | | ✓ | | ✓ | | ✓ | | ✓ | | ✓ | | | ✓ |
| Current stress severity (PSS-10) | | ✓ | | | | ✓ | | | | ✓ | | | | | | ✓ |
| Chronic Stress ratings | | ✓ | | | | ✓ | | | | ✓ | | | | | | ✓ |
| Functioning in stress and depression | | ✓ | | | | ✓ | | | | ✓ | | | | | | ✓ |
| Stressor inventory | | ✓ | | | | ✓ | | | | ✓ | | | | | | ✓ |
| Participant experience | | ✓ | | | | ✓ | | | | | | | | | | ✓ |
| Video journal | | ✓ | ✓ | ✓ | ✓ | ✓ | ✓ | ✓ | ✓ | ✓ | ✓ | ✓ | ✓ | ✓ | | |
| Weekly symptom survey | | ✓ | ✓ | ✓ | ✓ | ✓ | ✓ | ✓ | ✓ | ✓ | ✓ | ✓ | ✓ | ✓ | | ✓ |
| Daily mood watch survey (4x/day) | | Daily | | | | | | | | | | | | | ✓ | |
| Mood timestamps on watch (ad hoc) | | | | | | | | | | | | | | | ✓ | |
| Watch sensors | | | | | | | | | | | | | | | ✓ | |
| Beddit sensors | | | | | | | | | | | | | | | ✓ | |

***Supplementary Figure 11*** Assessment schedule for the DMHS-P2 Study. The exit session was scheduled approximately 12 weeks after the intake, but no later than 98 days after the intake. [a] For times when the exit session occurred 13-14 weeks after the intake, participants continued to receive Daily Mood Watch Surveys to complete four times daily until the exit session was completed. **Abbreviations:** Holmes-Rahe: Holmes-Rahe Life Stress Inventory; PSQI: Pittsburgh Sleep Quality Index; Maslach Burnout: Maslach Burnout Inventory; SAGE-SR = Screening Assessment for Guiding Evaluation-Self-Report; MADRS: Montgomery-Åsberg Depression Rating Scale; HAM-A: Hamilton Anxiety Rating Scale; SCID: Structured Clinical Interview for DSM Disorders; CAT-MH: Computerized Adaptive Testing for Mental Health; PHQ-8: Patient Health Questionnaire-8; GAD-7: Generalized Anxiety Disorder 7-item scale; PSS-10: Perceived Stress Scale-10.

#### SUPPLEMENTARY TABLES

#### SUPPLEMENTARY TABLE 1

| DMHS Eligibility Criteria |
| --- |
| Inclusion:<br>• At least 18 years of age<br>• Fluent in English<br>• Own a functioning iOS smartphone (iPhone 8 or newer, with iOS 15 or newer) with access to data plan and WiFi*<br>• Willingness to use issued Apple Watch for the duration of the study<br>• Confirmed status as a UCLA Health System patient with willingness to provide HIPAA Authorization for access to electronic health record or confirmed status as a matriculating UCLA student and willingness to provide access to student academic records<br>• Reside in the US for the duration of the study<br>• Able to read and understand a written informed consent form<br>• Willingness to participate in study assessments and use provided devices |
| Exclusion:<br>• Any self-reported diagnosis of major neurological condition impairing mobility, cognition, or language ability including multiple sclerosis, Parkinson disease or other movement disorder, motor neuron disease, stroke, or dementia<br>• Any self-reported other diagnosis involving chronic mobility impairment including spinal cord injuries, or severe osteoarthritis of knee or hip<br>• Self-reported diagnosis of schizophrenia<br>• Self-reported latex allergy<br>• Self-reported scars or tattoos covering the top sides of both wrists<br>• Self-reported blindness or visual impairment that is not correctable with glasses/contact lenses<br>• Previous participation in either of the two pilot studies (DMHS-P1, DMHS-P2) where the participant did not complete a certain number of assessments |

***Supplementary Table 1*** Digital Mental Health Study (DMHS) Eligibility Criteria. *Some participants who were otherwise eligible but did not have an iPhone were offered a loaner iPhone for use during the study. These individuals were responsible for their own data plan and were required to return the loaner iPhone at the end of their study participation.

#### SUPPLEMENTARY TABLE 2

| Type | Instrument Short Name | Domain | Description | Method of Administration | Frequency |
|---|---|---|---|---|---|
| Mental health assessments | PHQ-9 | Depression symptoms | Patient Health Questionnaire-9: Measure severity of current depression symptoms, including suicidal ideation | Self-report | Quarterly |
| | PHQ-8 | Depression symptoms | Patient Health Questionnaire-8: Measure severity of current depression symptoms, without item on suicidal ideation | Self-report | Biweekly |
| | GAD-7 | Anxiety symptoms | Generalized Anxiety Disorder 7-item scale: Measure severity of current anxiety symptoms | Self-report | Biweekly |
| | PVSS | Anhedonia | Positive Valence Systems Survey: 21 item survey measuring response to a wide range of rewards | Self-report | 2x |
| | Monthly SI | Suicidal ideation | Customized version of the Columbia-Suicide Severity Rating Scale | Self-report | Monthly |
| | MADRS | Depression symptoms | Montgomery-Åsberg depression rating scale (MADRS). Administered by trained clinical rater for current depression symptom severity. | Clinical interview | 2x |
| | YMRS | Mania | Young Mania Rating Scale. Administered by trained clinical rater for current hypomania or mania symptom severity. | Clinical interview | 2x |
| | CAT-MH | Mental Health Symptoms | Computerized Adaptive Test for Mental Health: Administered to assess current depression, anxiety, mania, suicidal ideation (SI) symptoms, post-traumatic stress disorder, and substance use disorders | Self-report | 4x |
| | SAGE-SR | Diagnostic screening | The Screening Assessment for Guiding Evaluation: Self Report. Administered for a comprehensive diagnostic history | Self-report | 2x |
| | SCID | Diagnosis | Structured Clinical Interview for the DSM-5: Computerized version via NetSCID administered by trained clinical rater for diagnoses of major mental disorders. | Clinical interview | 1x |
| Mood EMA | Daily Reflection | Mood - Daily lookback | Custom survey to describe your state over the last 24 hours: happy, sad, stressed, calm, anxious, energetic. Administered after 4pm daily | Self-report | Daily |
| | Current Mood EMA | Mood - Momentary | Custom survey asking how happy, sad, irritable, anxious, energetic you feel right now. Administered four times per day for one week straight to measure diurnal variation in mood | Self-report | Quarterly burst |
| | Energy & Mood | Mood - Momentary | Custom survey to measure affective valence and vigor in the moment. Administered with Video Journal | Self-report | Monthly |
| | Video Journal | Mood - Expressive | Self-guided, user-initiated AV recording of response to three mood-related questions | Self-report | Monthly |
| | Stress logs on Watch | Stress - Momentary | Time stamps of stress level made using Apple Watch. Optional | Self-report | Daily |
| Well being | Well-being | Mental well-being | Brief survey of sense of well-being, social relationships, and life purpose | Self-report | Monthly |
| | PSS-10 | Stress perception | Perceived Stress Scale: Measure perception of recent stress | Self-report | Monthly |

| Type | Instrument Short Name | Domain | Description | Method of Administration | Frequency |
|---|---|---|---|---|---|
| | Monthly symptoms and events | Symptoms - other | Custom survey to assess additional symptoms and life events in the past month, covering constructs not collected in the other surveys | Self-report | Monthly |
| | Maslach | Burnout | Maslach Burnout Survey: 22-item scale measuring workplace and occupational burnout | Self-report | 2x |
| Risk factors and covariates | Demographics | Characteristics | Custom self-report measure of demographics to capture age, sex at birth, race, ethnicity, education, socioeconomic status, and for UCLA student participants, student status | Self-report | 1x |
| | Medical comorbidities | Medical History | National Network of Depression Centers Comorbidity Questionnaire: Assessment of 15 medical conditions that are commonly comorbid with depression | Self-report | 2x |
| | Treatment History | Medical History | Current health assessments including medication history and treatment history | Self-report | 2x |
| | Current Medications | Medical History | Current medication use collected via interview with study staff | Clinical interview | 2x |
| | Early Adversity & Parental Bonding | Stressful Events - Early | Assess parental bonding and exposure to early abuse. Adapted from Parental Bonding Instrument and Childhood Trauma Questionnaire. | Self-report | 1x |
| | Lifetime stressful events | Stressful Events - Lifetime | Assess exposure to major stressful life events. Adapted from List of Threatening Experiences. | Self-report | 2x |
| | IPIP-NEO | Trait Neuroticism | International Personality Item Pool - Neuroticism, Extraversion, and Openness. 24 item measure to assess trait neuroticism | Self-report | 2x |
| | CD-RISC | Resilience | Connor-Davidson Resilience Scale. 25-item measure of stress coping ability | Self-report | 2x |
| | Peripartum | Pregnancy and childbirth | Custom survey administered to women to assess history of pregnancy and childbirth | Self-report | 2x |
| | PSQI | Sleep quality | Pittsburgh Sleep Quality Index: Assess recent sleep quality | Self-report | 2x |
| | Routines | Daily activities | Custom survey to assess daily activities and phone usage | Self-report | 3x |
| | Holmes-Rahe | Stressful Events - Recent | Holmes-Rahe Life Stress Survey: Assess recent exposure to moderate-to-major stressful life events | Self-report | quarterly |
| | Hair sample survey | Cortisol | Custom survey about hair color, hair care routines, and medication use that may affect hair cortisol results, administered only in participants providing hair samples | Self-report | 2x |
| Other | Participant experience | Study satisfaction | User experience questionnaires. Custom questionnaires to assess participant experience in the research study | Self-report | 4x |

***Supplementary Table 2*** Assessments used in the DMHS. [a] See Supplementary Table 3 for more information about the *Monthly Symptoms and Events* survey. [b] See *Supplementary Table 4A* for more information about Daily Mood surveys and EMAs in the DMHS. **Abbreviations:** PHQ-9: Patient Health Questionnaire-9; PHQ-8: Patient Health Questionnaire-8; GAD-7: Generalized Anxiety Disorder 7-item scale; PSS-10: Perceived Stress Scale-10; PVSS: Positive Valence Systems Scale; MADRS: Montgomery-Åsberg Depression Rating Scale; YMRS: Young Mania Rating Scale; CAT-MH: Computerized Adaptive Testing for Mental Health; SAGE-SR = Screening Assessment for Guiding Evaluation-Self-Report; SCID: Structured Clinical Interview for DSM Disorders; IPIP: International Personality Item Pool Neuroticism scale; CD-RISC: Connor-Davidson Resilience Scale; PSQI: Pittsburgh Sleep Quality Index; Holmes-Rahe: Holmes-Rahe Life Stress Inventory.

#### SUPPLEMENTARY TABLE 3

| If this PHQ item is endorsed... | ... these contingent follow-up questions are prompted | Response options |
|---|---|---|
| 1. Anhedonia | How often did you engage in fewer activities due to loss of interest or enjoyment? | • Not at all<br>• Several days<br>• More than half the days<br>• Nearly every day |
| 3. Sleep disturbance | Over the last 2 weeks, how often have you been bothered by any of the following problems?<br>• Trouble falling asleep<br>• Trouble staying asleep<br>• Sleeping too much | |
| 5. Appetite | Over the last 2 weeks, how often have you been bothered by any of the following problems?<br>• Poor appetite<br>• Overeating | |
| 8. Psychomotor | Over the last 2 weeks, how often have you been bothered by any of the following problems?<br>• Moving slowly<br>• Speaking slowly<br>• Being so fidgety or restless that you have been moving around a lot more than usual | |

**Supplementary Table 3A**. Contingent questions appended to the PHQ-8 and PHQ-9. If the participant endorsed the question listed in Column 1 with 'Several Days' or more, the corresponding contingent question(s) would be administered at the end of the survey, immediately following completion of all standard 8 or 9 questions.

<table>
<tr><th colspan="2">Over the last 2 weeks, how often have you experienced the following?</th></tr>
<tr><td>Sudden, intense mood swings</td><td rowspan="11">• Not at all<br>• Several days<br>• More than half the days<br>• Nearly every day</td></tr>
<tr><td>When other people have strong emotional responses, I don't feel anything at all</td></tr>
<tr><td>Trouble with decision-making</td></tr>
<tr><td>Feel lonely</td></tr>
<tr><td>Rapid heartbeat or heart palpitations (racing, pounding, fluttering, or skipping a beat)</td></tr>
<tr><td>Decreased sexual drive or interest compared to last month</td></tr>
<tr><td>Muscle tension, headaches, neck or back pain</td></tr>
<tr><td>Other physical pain not covered by the last question</td></tr>
<tr><td>Avoid activities which might cause you embarrassment or in which you need to speak to people</td></tr>
<tr><td>Increased use of caffeine, tobacco, alcohol or other non-prescription substances compared to last month</td></tr>
<tr><td>Staying in (e.g., spent most of your time at home or dorm)</td></tr>
<tr><td>Weight change compared to last month</td><td>• Gained a lot of weight<br>• Gained a little weight<br>• Weight has not changed<br>• Lost a little weight<br>• Lost a lot of weight</td></tr>
<tr><td>Was this weight change desired?</td><td>• Yes<br>• No</td></tr>
<tr><td>Did you have physical injury or illness which resulted in decreased activity?</td><td>• Yes<br>• No<br>• N/A</td></tr>
<tr><td>First day of last period</td><td>•</td></tr>
<tr><td>Are you currently pregnant?</td><td>• Yes<br>• No<br>• N/A</td></tr>
</table>

***Supplementary Table 3B*** The *Monthly Symptoms and Events* survey was a custom survey consisting of 13 questions, some of which had follow-up queries, which were assessed every four weeks throughout the DMHS. The survey was designed to cover mental constructs related to mental health that were not adequately assessed in off-the-shelf assessment instruments.

#### SUPPLEMENTARY TABLE 4

| EMA | Frequency | Questions | Response Options | Example |
|---|---|---|---|---|
| DMHS<br>Daily Reflection EMA | Daily at:<br>• 4 pm (PST) | Please select the answer to each question that best describes your state over the last 24 hours.<br>• Stress<br>• Happy<br>• Anxious<br>• Calm<br>• Sad<br>• Energetic | 0 – Very slightly or not at all<br>1 – A little<br>2 – Moderately<br>3 – Quite a bit<br>4 – Extremely | 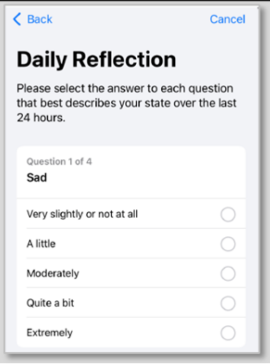 |
| | | Overall, how have you felt today? | -3 – Very unpleasant<br>-2 – Moderately unpleasant<br>-1 – A little unpleasant<br>0 – Neutral<br>1 – A little pleasant<br>2 – Moderately pleasant<br>3 – Very pleasant | |
| DMHS<br>High Frequency<br>Current Mood EMA | Completed three times each day for one week each quarter, choosing 3 of the 4 time options below:<br>• 6 am (PST)<br>• 12 pm (PST)<br>• 6 pm (PST)<br>• 12 am (PST) | • How sad do you feel right now?<br>• How irritable do you feel right now?<br>• How anxious do you feel right now?<br>• How energetic do you feel right now?<br>• How happy do you feel right now? | 0 – Very little<br>to<br>7 – Very much | 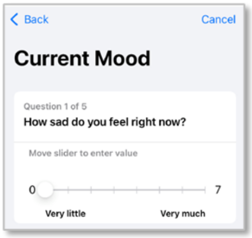 |

***Supplementary Table 4A.*** Ecological Momentary Assessment (EMA) in the DMHS. In the 52-week schedule, High Frequency Current Mood EMAs were scheduled during weeks 3, 15, 27, and 40. The High Frequency Current Mood Survey was made available at 4 times each day to provide participants with variable schedules the opportunity to complete three surveys each day.

| Task | Frequency | Questions |
|---|---|---|
| Video Journal, DMHS | Monthly | 1. What has been on your mind recently?<br>2. Did you feel angry, stressed, or sad this week? If so, why?<br>3. Describe the best thing that happened this week. |

***Supplementary Table 4B.*** Video Journal prompts in the DMHS.

| Session | Questions |
|---|---|
| **Check-in 1** | 1. What questions do you have about the study?<br>2. What is your occupation?<br>3. Can you say a little more about what you do?<br>4. What are your activities like during the day?<br>5. Do you have any pets? Did you ever have any pets? Tell me about them.<br>6. How is your week going?<br>7. What plans do you have for the weekend?<br>8. Tell me about one of your favorite vacations. Where did you go?<br>9. What did you like about it? What didn't work out?<br>10. What got your attention in the news recently? |

| | |
|---|---|
| | 11. What did you do this week that you enjoyed? |
| **Check-in 2** | 1. What questions do you have about the study?<br>2. What is the best thing about the study? What do you like about it?<br>3. What is the worst thing about the study? What don't you like about it?<br>4. What would make the study better?<br>5. How is your week going?<br>6. What have you been working on recently?<br>7. How are your weekend plans shaping up?<br>8. Tell me about your most recent vacation. Where did you go?<br>9. What did you like about it? What didn't work out?<br>10. What got your attention in the news recently?<br>11. What did you do that you enjoyed since we last spoke? |
| **SCID** | 1. What questions do you have about the study?<br>2. What was the best thing about the study? What did you like about it?<br>3. What was the worst thing about the study? What didn't you like about it?<br>4. Which self-report did you feel most comfortable with? (watch, phone surveys, video journal)?<br>5. How is your week going?<br>6. How are your weekend plans shaping up?<br>7. What is the next trip you have planned? Can you tell me more about it?<br>8. Do you have any plans for the holidays / What did you do for the holidays this year?<br>9. Are you excited about the rest of 2021?<br>10. If you could wave a magic wand and change on thing, what would it be?<br>11. What got your attention in the news recently?<br>12. Describe a happy event that occurred since we last spoke |

***Supplementary Table 4C.*** Rater prompts for the 10-minute conversation prior to the clinical interview.

| | |
|---|---|
| Below are 4 statements related to your wellbeing. Please respond how you feel about each of these statements, reflecting on the past month. | |
| I feel positive, happy, content | 0, Disagree<br>1, Slightly disagree<br>2, Mixed or neither agree nor disagree<br>3, Slightly agree<br>4, Agree |
| My social relationships are supportive and rewarding[a] | |
| I lead a purposeful and meaningful life[a] | |
| I listen for information from my body about my emotional state[b] | |

***Supplementary Table 4D.*** The Well-being scale was a customized survey that included selected questions from two other scales. [a] The Flourishing Scale (Deiner at al. 2009) [2]; [b] Multidimensional Assessment of Interoceptive Awareness (MAIA) (Mehling et al., 2012) [3]

#### SUPPLEMENTARY TABLE 5

| Passive data types collected from device sensors in DMHS | |
|---|---|
| **A. HealthKit Data** | |
| **Category** | **Variables Collected** |
| Cardiovascular | HeartRate, HeartbeatSeries, RestingHeartRate, WalkingHeartRateAverage, HeartRateVariabilitySDNN, LowHeartRateEvent, HighHeartRateEvent, IrregularHeartRhythmEvent, Electrocardiogram |
| Cardiofitness | VO2Max, HeartRateRecoveryOneMinute, LowCardioFitnessEvent,OxygenSaturation, CardioFitnessMedicationsUse, PeripheralPerfusionIndex, HeartRateRecoveryOneMinute, LowCardioFitnessEvent, SixMinuteWalkTestDistance |
| Activity | StepCount, DistanceWalkingRunning, AppleStandTime, AppleStandHour, StairAscentSpeed, StairDescentSpeed, WalkingSpeed, WalkingStepLength BasalEnergyBurned, ActiveEnergyBurned, FlightsClimbed, AppleExerciseTime, PhysicalEffort, EstimatedWorkoutEffortScore, WorkoutEffortScore, RunningStrideLength, RunningVerticalOscillation, RunningGroundContactTime, RunningPower, RunningSpeed, CyclingPower, CyclingFunctionalThresholdPower, CyclingSpeed,CyclingCadence, DistanceCrossCountrySkiing, CrossCountrySkiingSpeed, DistanceRowing, RowingSpeed, DistanceSkatingSports, DistancePaddleSports, PaddleSportsSpeed, DistanceCycling, PushCount,DistanceSwimming, SwimmingStrokeCount, DistanceDownhillSnowSports |
| Mobility quality | WalkingDoubleSupportPercentage, WalkingAsymmetryPercentage |
| Sleep | SleepSchedule, SleepDurationGoal, SleepAnalysis |
| Hearing | Audiogram, EnvironmentalAudioExposure, EnvironmentalAudioExposureEvent, HeadphoneAudioExposure, HeadphoneAudioExposureEvent |
| Body measurements | BodyMass |
| State of mind and mindfulness | StateOfMind, MindfulSession |
| Light Exposure | TimeInDaylight, UVExposure |
| Medical Records | AllergyRecord, ConditionRecord, ImmunizationRecord, LabResultRecord, MedicationRecord, ProcedureRecord, VitalSignRecord |
| Blood Pressure [a] | BloodPressureSystolic, BloodPressureDiastolic |
| Medical Symptoms [a] | User stated occurrences of AbdominalCramps, Bloating,Constipation, Diarrhea, Heartburn, Nausea, Vomiting, AppetiteChangesChills, Dizziness,Fainting, Acne, BladderIncontinence, BreastPain, ChestTightnessOrPain, Coughing, DrySkin, Fatigue, Fever, HairLoss, Headache, HotFlashes, LowerBackPain, MemoryLapse, MoodChanges, NightSweats, PelvicPain, RapidPoundingOrFlutteringHeartbeat, RunnyNose, ShortnessOfBreath, SleepChanges, SinusCongestion, SkippedHeartbeat, SoreThroat,Wheezing |
| Reproductive Health [a] | User stated occurrences of MenstrualFlow, IntermenstrualBleeding, IrregularMenstrualCycles, InfrequentMenstrualCycles, PersistentIntermenstrualBleeding, ProlongedMenstrualPeriods, CervicalMucusQuality, Contraceptive, Lactation, OvulationTestResult, Pregnancy, SexualActivity, VaginalDryness, BasalBodyTemperature |

| B. SensorKit Data | |
|---|---|
| **Category** | **Variables Collected** |
| Accelerometer | High fidelity acceleration data from iPhone and Apple Watch in x,y,z directions. |
| AmbientLightSensor | Lux, chromaticity when screen is on |
| DeviceUsageReport | Number of unlocks, number of screen wakes, total duration unlocked, usage time of apps by category, number of notifications by category, usage time of web domains by category. Aggregated over 15-minute time periods. |
| FaceMetrics | Collected after unlock, and during messaging app usage (No images or video was collected, and no face recognition data was collected.)<br>ARFaceAnchor: 52 Blendshapes such as eyeBlinkLeft, head rotation and translation, gaze direction (face geometry was not collected)<br>Whole Face Expressions: BrowFurrowAndEyesWideAndMouthPressedTight, Baseline, NoseWrinkleOrUpperLipRaise, BrowRaiseWithFurrowAndEyeWidenAndMouthStretch, LipRaiseAndCheekRaise, InnerBrowRaiseAndMouthCornerDepress, BrowRaiseWithoutFurrowAndEyeWidenAndJawDrop.<br>PartialFaceExpressions: InnerBrowRaise, BrowRaiseWithFurrow, BrowRaiseWithoutFurrow, BrowFurrow, MouthCornerDepress, MouthStretch, MouthPressedOrTight, NoseWrinkle |
| Gyroscope | High fidelity rotation data from iPhone and Apple Watch (x, y, z) |
| KeyboardMetrics | Collected during keyboard bringups. (The words typed were not collected.)<br>Productivity: totalWords, totalTaps, totalTypingDuration, number of typing episodes, totalDrags, totalEmojis.<br>Kinematics: Typing speed, Total typing pauses, hold time, flight time, interkey delay.<br>Corrections: totalAlteredWords, totalDeletes, totalAutoCorrections.<br>Tap position error: upErrorDistance, downErrorDistance.<br>Paths: totalPaths, totalPathDuration, totalPathLength, path typing speed, total mid-path pauses, path error distance ratio.<br>Sentiment: total_positive_words, total_down_words, total_anxiety_words, total_anger_words, total_health_feeling_words, total_death_words, total_absolutist_words. Emoji categories: total_positive_emojis, total_sad_emojis, total_anxiety_emojis, total_anger_emojis, total_health_feeling_emojis, total_low_energy_emojis, total_confused_emojis |
| MessagesUsageReport | (Excludes any message content and information about people messaged.)<br>Total outgoing messages, total incoming messages, total unique contacts. Aggregated over 30-minute time periods. |
| PedometerData | CMPedometerData: current_pace, current_cadence, avg_active_pace, cumulative stepcount, cumulative distance, cumulative flights ascended, cumulative flights descended. |
| PhoneUsageReport | Total outgoing calls, total incoming calls, total unique contacts, total call duration. Aggregated over 24-hour periods. |
| SpeechMetrics | Collected during phone calls and VOIP. (No audio or conversation content was collected.)<br>SFSpeechRecognitionMetadata: speakingRate, speechDuration, averagePauseDuration, SpeechStartTimestamp, jitter, shimmer, pitch, voicing.<br>Confidence value of transcription, but not the transcription itself.<br>SpeechExpression: valence, activation, dominance, mood.<br>SoundClassification: laughter, shouting, isSpeech (boolean) |

| | |
|---|---|
| Visits[b] | Location category (home, work, other) [b], distance from home, de-identified location ID, approximate arrival and departure times. (Raw GPS was not collected) |
| WristDetection | Times when Apple Watch is worn. |

***Supplementary Table 5*** Digital sensing metrics in the DMHS. [a] Available when logged by the participant; [b] Raw GPS or specific location was not collected; instead, time spent in frequently visited locations was collected using an anonymous identifier.

#### SUPPLEMENTARY TABLE 6

| Data Type | Requirements by Tier | | | Data Missingness Reach Out |
|---|---|---|---|---|
| | Base | Silver | Gold | |
| Assessment Visits (Intake, Week 10 Check-in, Week 46 Check-in, Exit) | Intake & Week 10 | Intake & Week 10 | Intake & Week 10 | Up to 3 contact attempts to schedule each assessment visit; Flat rate compensation awarded for each assessment visit |
| Daily reflection survey | 4/7 days/week | 5/7 days/week | 6/7 days/week | Not completed within 3 most recent consecutive days since activation |
| Current Mood EMA (High frequency burst) | 4/7 days | 5/7 days | 6/7 days | Not completed within 2 most recent consecutive days |
| Other Tasks in Study App: PHQ8, GAD7, PSS10, Monthly Symptoms, Energy & Mood, Video Journals | 58% per week | 75% per week | 85% per week | Not completed within 7 days of activation |
| Surveys via emailed link (PHQ9, SI, CAT-MH, SAGE-SR, PVSS, Well-being, Maslach) | 58% per week | 75% per week | 85% per week | Not completed within 14 days of activation |
| Stress logs on Watch | 2/week | 3/week | 4/week | No reach out, compensate as bonus |
| Watch wear (wearing ≥ 20 hr/day) | 4/7 days/week | 5/7 days/week | 6/7 days/week | Threshold not met for 3 consecutive days |
| Beddit data | no min | no min | no min | No data for 3 consecutive days |
| HealthKit data | authorized | authorized | authorized | If Stand Hours data missing for 3 consecutive days |
| SensorKit data | authorized | authorized | authorized | If frequently Visited Locations type has 0 visits logged over 7 consecutive days |
| Indicator that Watch app, sleep schedule, or Bedit app not installed or deleted; | Installed | Installed | Installed | As soon as indicated in compliance report |
| Indicator of data received from multiple phones | one phone | one phone | one phone | As soon as indicated in compliance report |

***Supplementary Table 6*** Compliance requirements for DMHS. Base compliance was required to remain in the study, while Silver and Gold received extra compensation. Data missingness reach out thresholds shown for each data type.

### SUPPLEMENTARY TABLE 7

**Completion rates for selfreport assessments in the Digital Mental Health Study**

| | BASELINE | MONTH 1 | MONTH 2 | MONTH 3 | MONTH 4 | MONTH 5 | MONTH 6 | MONTH 7 | MONTH 8 | MONTH 9 | MONTH 10 | MONTH 11 | MONTH 12 | MONTH 13 | EXIT |
|---|---|---|---|---|---|---|---|---|---|---|---|---|---|---|---|
| Demographics | **3332** *100%* | | | | | | | | | | | | | | |
| Medical comorbidities | **3332** *100%* | | | | | | | | | | | | | | **3332** *100%* |
| Treatment history | **3332** *100%* | | | | | | | | | | | | | | **3332** *100%* |
| Early adversity & parental bonding | **3332** *100%* | | | | | | | | | | | | | | |
| Lifetime stressful events | **3332** *100%* | | | | | | | | | | | | | | **3332** *100%* |
| Recent stressful events (Holmes-Rahe) | **3332** *100%* | | | **2848** *85%* | | | **2814** *84%* | | | **2735** *82%* | | | | **2684** *81%* | **3332** *100%* |
| Trait neuroticism | **3332** *100%* | | | | | | | | | | | | | | **3332** *100%* |
| Resilience | **3332** *100%* | | | | | | | | | | | | | | **3332** *100%* |
| Routines | **3332** *100%* | | | | | | **2824** *85%* | | | | | | | | **3332** *100%* |
| Peripartum (females only) | **1908** *57%* | | | | | | | | | | | | | | **1910** *57%* |
| Medication Use | **3332** *100%* | | | | | | | | | | | | | | **3332** *100%* |
| Clinical psychiatric screener (SAGE-SR) | | | **2733** *82%* | | | | | **2712** *81%* | | | | | | | |
| Depression rating scale (MADRS) | | | | **3332** *100%* | | | | | | | | | **3332** *100%* | | |
| Mania rating scale (YMRS) | | | | **137** *4%* | | | | | | | | | **112** *3%* | | |
| Diagnostic interview (Net-SCID) | | | | | | | | | | | **236** *7%* | | | | |
| Current mental health symptoms (CAT-MH) | | | **2704** *81%* | | | | | **2662** *80%* | | | | | | | **3323** *100%* |
| Current depression severity with SI (PHQ-9) | **3331** *100%* | | | **3328** *99%* | | | | **2848** *85%* | | | **236** *7%* | | **3032** *91%* | | **3331** *100%* |
| Monthly SI | | **3049** *92%* | **2954** *89%* | | **2827** *85%* | **2836** *85%* | **2811** *84%* | | **2740** *82%* | **2732** *82%* | **2734** *82%* | **2690** *81%* | | **2674** *80%* | |
| Well-being | | **3043** *91%* | **2947** *88%* | **2846** *85%* | **2824** *85%* | **2835** *85%* | **2810** *84%* | **2850** *86%* | **2734** *82%* | **2728** *82%* | **2728** *82%* | **2686** *81%* | **2667** *80%* | **2672** *80%* | **3332** *100%* |
| Sleep quality (PSQI) | | | **2942** *88%* | | | | | | | | | | | | **3332** *100%* |
| Burnout (Maslach) | | | | | | | **2804** *84%* | | | | | | | | **3332** *100%* |
| Anhedonia (PVSS) | | | | | | | **2795** *84%* | | | | | | | | **3332** *100%* |
| Current stress severity (PSS-10) | **3303** *99%* | **3253** *98%* | **3208** *96%* | **3175** *95%* | **3128** *94%* | **3101** *93%* | **3030** *91%* | **2975** *89%* | **2884** *87%* | **2898** *87%* | **2836** *85%* | **2800** *84%* | **2789** *84%* | **2555** *77%* | **3332** *100%* |
| Monthly life events and additional symptoms | **3303** *99%* | **3248** *97%* | **3211** *96%* | **3171** *95%* | **3130** *94%* | **3093** *93%* | **3022** *91%* | **2951** *89%* | **2874** *86%* | **2885** *87%* | **2808** *84%* | **2789** *84%* | **2771** *83%* | **2540** *76%* | **3332** *100%* |
| Participant experience | **3300** *99%* | | | | | | | **3041** *91%* | | | | | | | **3332** *100%* |
| Video journal | **3269** *98%* | | **3173** *95%* | **3054** *92%* | **3018** *91%* | **2854** *86%* | **2699** *81%* | **2695** *81%* | **2601** *78%* | **2469** *74%* | **2393** *72%* | **2383** *72%* | **2315** *69%* | **2292** *69%* | |
| Energy & mood | **3310** *99%* | | **3285** *99%* | **3224** *97%* | **3204** *96%* | **3126** *94%* | **3052** *92%* | **3029** *91%* | **2940** *88%* | **2835** *85%* | **2784** *84%* | **2786** *84%* | **2731** *82%* | **2728** *82%* | |

**Bi-Weekly Tasks**

| | BASELINE | WEEK 2 | WEEK 4 | WEEK 6 | WEEK 8 | WEEK 10 | WEEK 12 | WEEK 14 | WEEK 16 | WEEK 18 | WEEK 20 | WEEK 22 | WEEK 24 | WEEK 26 | WEEK 28 |
|---|---|---|---|---|---|---|---|---|---|---|---|---|---|---|---|
| PHQ-8 | **3304** *99%* | **3278** *98%* | **3269** *98%* | **3252** *98%* | **3233** *97%* | **3209** *96%* | **3183** *96%* | **3191** *96%* | **3162** *95%* | **3128** *94%* | **3118** *94%* | **3079** *92%* | **3051** *92%* | **3016** *91%* | **2991** *90%* |
| GAD-7 | **3304** *99%* | **3272** *98%* | **3259** *98%* | **3244** *97%* | **3220** *97%* | **3193** *96%* | **3162** *95%* | **3181** *95%* | **3147** *94%* | **3114** *93%* | **3096** *93%* | **3057** *92%* | **3019** *91%* | **2987** *90%* | **2961** *89%* |
| | | | WEEK 30 | WEEK 32 | WEEK 34 | WEEK 36 | WEEK 38 | WEEK 40 | WEEK 42 | WEEK 44 | WEEK 46 | WEEK 48 | WEEK 50 | WEEK 52 | EXIT |
| PHQ-8 | | | **2950** *89%* | **2909** *87%* | **2874** *86%* | **2846** *85%* | **2871** *86%* | **2843** *85%* | **2770** *83%* | **2799** *84%* | **2778** *83%* | **2775** *83%* | **2782** *83%* | **2686** *81%* | **3330** *99%* |
| GAD-7 | | | **2911** *87%* | **2866** *86%* | **2842** *85%* | **2818** *85%* | **2849** *86%* | **2818** *85%* | **2738** *82%* | **2756** *83%* | **2742** *82%* | **2736** *82%* | **2736** *82%* | **2640** *79%* | **3332** *100%* |

***Supplementary Table 7*** Completion rates of self-report assessments administered in the DMHS among those who participated in the entire study. The denominator for all percentage calculations is the final 3,332 study completers. **Abbreviations:** Holmes-Rahe: Holmes-Rahe Life Stress Inventory; SAGE-SR = Screening Assessment for Guiding Evaluation-Self-Report; MADRS: Montgomery-Åsberg Depression Rating Scale; YMRS: Young Mania Rating Scale; SCID: Structured Clinical Interview for DSM Disorders; CAT-MH: Computerized Adaptive Testing for Mental Health; PHQ-9: Patient Health Questionnaire-9; Pittsburgh Sleep Quality Index; Maslach Burnout: Maslach Burnout Inventory; PVSS:

Positive Valence Systems Scale; PSS-10: Perceived Stress Scale-10; PHQ-8: Patient Health Questionnaire-8; GAD-7: Generalized Anxiety Disorder 7-item scale.

#### SUPPLEMENTARY TABLE 8

| **DMHS-P1 Eligibility Criteria** |
| --- |
| Inclusion:<br>• At least 18 years of age<br>• Fluent in English<br>• Own functioning iOS smartphone (iPhone 7 or newer, with iOS 13.5.1 or newer) with access to data plan and WiFi<br>• Willingness to use issued Apple Watch for the duration of the study<br>• Confirmed status as UCLA Health System patient, willingness to provide HIPAA Authorization for access to medical record<br>• Reside in the US for the duration of the study<br>• Able to read and understand a written informed consent form<br>• Willingness to participate in study assessments and use provided devices<br>Exclusion:<br>• Any self-reported diagnosis of major neurological condition impairing mobility, cognition, or language ability including multiple sclerosis, Parkinson disease or other movement disorder, motor neuron disease, stroke, or dementia<br>• Any self-reported other diagnosis involving chronic mobility impairment including spinal cord injuries, or severe osteoarthritis of knee or hip<br>• Self-reported diagnosis of schizophrenia<br>• Any self-reported facial muscle impairment of significant facial scarring, and no Botox within the last 6 months<br>• No planned travel outside of the US during the duration of the study |

***Supplementary Table 8*** DMHS-P1 Eligibility Criteria.

#### Supplementary Table 9

| **DMHS-P2 Eligibility Criteria** |
|---|
| Inclusion:<br>• At least 18 years of age<br>• Fluent in English<br>• Own functioning iOS smartphone (iPhone 7 or newer, with iOS 14 or newer) with access to data plan and WiFi<br>• Willingness to use issued Apple Watch for the duration of the study<br>• Confirmed status as UCLA Health System patient, willingness to provide HIPAA Authorization for access to medical record<br>• Reside in the US for the duration of the study<br>• Able to read and understand a written informed consent form<br>• Willingness to participate in study assessments and use provided devices<br>Exclusion:<br>• Any self-reported diagnosis of major neurological condition impairing mobility, cognition, or language ability including multiple sclerosis, Parkinson disease or other movement disorder, motor neuron disease, stroke, or dementia<br>• Any self-reported other diagnosis involving chronic mobility impairment including spinal cord injuries, or severe osteoarthritis of knee or hip<br>• Self-reported diagnosis of schizophrenia<br>• Having previously participated in our first pilot study (DMHS-P1) |

***Supplementary Table 9*** DMHS-P2 Eligibility Criteria.

#### SUPPLEMENTARY TABLE 10

| EMA | Frequency | Questions | Response Options | Example |
|---|---|---|---|---|
| DMHS-P1<br>Daily Mood EMA | Daily at:<br>• 4 pm (PST) | *Low burden group* [a]:<br>Over the past 24 hours, how much were you:<br>• Energetic<br>• Happy<br>• Sad<br>• Stressed<br><br>*High burden group* [b]:<br>Over the past 24 hours, how much were you:<br>• Energetic<br>• Happy<br>• Sad<br>• Stressed<br>• Tired<br>• Little interest or pleasure in doing things<br>• Outgoing<br>• Calm<br>• Anxious<br>• Trouble concentrating on things | 0 – Very slightly or not at all<br>1 – A little<br>2 – Moderately<br>3 – Quite a bit<br>4 – Extremely | 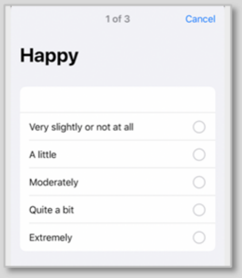 |

***Supplementary Table 10*** Ecological Momentary Assessment (EMA) in the DMHS-P1 study. [a] In DMHS-P1, participants were randomly assigned to either the Low (4 daily mood questions) or High Burden group (10 daily mood questions). [b] An example of the 10 daily mood questions presented to participants in the High Burden group; the selection of 10 daily mood questions was varied on each day of the week, with the items drawn from a larger set of 34 items presented to all participants at intake and exit.

#### SUPPLEMENTARY TABLE 11

| EMA | Frequency | Questions | Response Options | Example |
|---|---|---|---|---|
| DMHS-P2<br>High Frequency EMA | Four times daily [a] at:<br>• 8 am (PST)<br>• 12 pm (PST)<br>• 4 pm (PST)<br>• 8 pm (PST) | • How are you? | 0 – Not so great<br>1 – Okay<br>2 – Great | 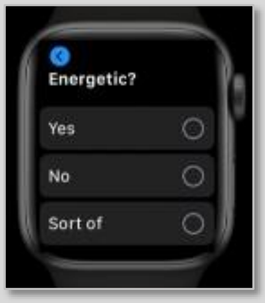 |
| | | • Are you feeling energetic right now? | 0 – No<br>1 – Sort of<br>2 - Yes | |
| | | • Are you feeling stressed right now? | 0 – No<br>1 – Sort of<br>2 - Yes | |
| | | • Well rested last night? [b] | 0 – No<br>1 – Sort of<br>2 - Yes | |

***Supplementary Table 11*** Ecological Momentary Assessment (EMA) in the DMHS-P2 study. [a] Four times daily mood ratings were made via Apple Research app installed on Apple Watch in DMHS-P2 Study. [b] Perceptions of sleep quality were only included in the first EMA set of each day.
